# Genome-wide analysis of longitudinal lung function and gas transfer in individuals with idiopathic pulmonary fibrosis

**DOI:** 10.1101/2022.03.28.22272832

**Authors:** Richard J Allen, Justin M Oldham, David A Jenkins, Olivia C Leavy, Beatriz Guillen-Guio, Carl A Melbourne, Shwu-Fan Ma, Jonathan Jou, John S Kim, CleanUP-IPF Investigators of the Pulmonary Trials Cooperative, William A Fahy, Eunice Oballa, Richard B Hubbard, Vidya Navaratnam, Rebecca Braybrooke, Gauri Saini, Katy M Roach, Martin D Tobin, Nik Hirani, Moira K B Whyte, Naftali Kaminski, Yingze Zhang, Fernando J Martinez, Angela L Linderholm, Ayodeji Adegunsoye, Mary E Strek, Toby M Maher, Philip L Molyneaux, Carlos Flores, Imre Noth, R Gisli Jenkins, Louise V Wain

## Abstract

**Background:** Idiopathic pulmonary fibrosis (IPF) is an incurable disease characterised by progressive scarring of the lungs. This leads to the lungs becoming stiffer, reducing lung capacity, and impeding gas transfer. We aimed to identify genetic variants associated with either declining lung capacity or gas transfer after diagnosis of IPF.

**Methods:** We performed a genome-wide meta-analysis of longitudinal measures of forced vital capacity (FVC) and diffusing capacity for lung of carbon monoxide (DLco) in individuals diagnosed with IPF from three studies. Suggestively significant variants were investigated further in an additional study. Variants were defined as significantly associated if they had a meta-analysis p<5×10^−8^, had consistent direction of effects across all studies and were nominally significant (p<0.05) in each study.

**Findings:** 1,048 individuals with measures of longitudinal FVC and 729 individuals with longitudinal measures of DLco passed quality control. In total, 4,560 measures of FVC and 2,795 measures of DLco and over 7 million genetic variants were included in the analysis. One variant located in an antisense RNA gene for Protein Kinase N2 (*PKN2*) showed a genome-wide significant association with FVC decline (−140 ml/year per risk allele, 95% CI [−180, −100], p=9.14×10^−12^).

**Interpretation:** These results identify a possible druggable target involved in promoting IPF disease progression.

**Funding:** Action for Pulmonary Fibrosis, Medical Research Council, Wellcome Trust, National Institute of Health/National Heart, Lung and Blood Institute

**Research in context:** *Evidence before this study:* Idiopathic pulmonary fibrosis (IPF) is a devastating disease where the lungs become scarred, this scarring leads to a reduced lung capacity, poorer rates of gas transfer and is eventually fatal. However, disease progression is highly variable and it is not clear why this is. To date, genome-wide association studies (GWAS) have identified 20 genetic loci associated with susceptibility to IPF. These genetic loci implicate genes involved with host defence, regulation of TGFβ signalling, telomere maintenance, cell-cell adhesion and spindle assembly as important biological processes involved in the pathogenesis of IPF. The GWAS variant with the strongest effect on disease risk is found in the promoter region of the *MUC5B* gene (rs35705950). Generally, the variants associated with IPF susceptibility show little or no association with disease progression, apart from the risk allele at rs35705950 which has been reported as having an association with improved survival times.

*Added value of this study:* Although genetic variants associated with disease risk have been widely studied, little has been reported to investigate the effect of genetics on progression of IPF. Here we present a GWAS of progressive IPF by identifying genetic variants associated longitudinal measures of lung health after diagnosis of IPF. We identify a genetic locus associated with a more rapid decline in lung capacity that lies in the RNA antisense gene of *PKN2*.

*Implications of all available evidence:* The novel genetic locus associated with a more rapid decline in lung capacity in individuals with IPF implicates a Rho/RAC effector protein. Effective treatments for IPF are desperately needed. There are currently PKN2 inhibitors under development meaning this analysis highlights a potential therapeutic target for IPF. We also show the genetic determinants of IPF progression appear to be distinct from those that drive IPF susceptibility.

## Introduction

Idiopathic pulmonary fibrosis (IPF) is a devastating lung disease characterised by an aberrant response to lung injury leading to the deposit of scar tissue in the lung alveoli. IPF has a prevalence of around 50 per 100,000 individuals and is more common in men, the elderly and those of European ancestry.

As the disease progresses, the scarring spreads throughout the lung. This scarring makes the lungs stiffer, making it more difficult to breathe and leading to reduced lung capacity. The scar tissue also impedes gas transfer in and out of the bloodstream. This decrease in lung health leads to a poorer quality of life and eventually death with half of individuals dying within 3 to 5 years of diagnosis. Two measures of lung health are commonly used to monitor disease progression in individuals with IPF. These are forced vital capacity (FVC, i.e. the total volume of air that can be expired measured through spirometry) and the diffusing capacity for lung of carbon monoxide (DLco, i.e. a measure of how much gas can pass between the air sacs and bloodstream).

Generally, both FVC and DLco decline over time in individuals with IPF. However, rates of lung function decline are highly variable between individuals with some individuals experiencing rapid decline and shorter survival times, while others experience relatively stable lung function and live for many years after diagnosis^1^.

There are many known genetic and environmental risk factors for IPF. Studies of sporadic and familial IPF have identified genetic risk factors that implicate host defence (such as mucus and pulmonary surfactant regulation), signalling (particularly regulation of TGFβ signalling), cell-cell adhesion (such as desmoplakin which plays a role in structural integrity of the epithelium), telomere maintenance (with IPF cases having shorter telomeres than age-matched controls), and spindle assembly as important processes in disease risk^2-7^.

Drug targets with supporting genetic evidence are twice as likely to be successful during drug development^8^. Little is known about genetic determinants of disease progression and identifying targets that modify disease process, rather than predisposing to disease, might yield therapeutic benefit.

We aimed to identify biological processes involved in disease progression by performing the first genome-wide association study (GWAS) of lung function and gas transfer decline in individuals with IPF.

## Methods

For this study we utilised a two-stage design. Firstly, variants from across the genome were tested for their association with longitudinal FVC and DLco separately in discovery GWASs. Variants showing evidence of being associated with rate of change of FVC or DLco in the discovery GWAS were then analysed in an independent analysis and variants reaching genome-wide significance (with support from all studies) were reported. Full summary statistics for the FVC and DLco genome-wide meta-analyses can be accessed from https://github.com/genomicsITER/PFgenetics.

### Study populations

For the discovery GWAS, we utilised longitudinal measures of FVC and DLco from IPF cases from three previously described studies (named as US, UK, UUS).

The US study^4^ (referred to as the “Chicago study” in some previous IPF GWAS) had three study centres that collected longitudinal data, namely the UC (University of Chicago), UPMC (University of Pittsburgh Medical Centre) and COMET (Correlating Outcomes with biochemical Markers to Estimate Time-progression) centres. The COMET study did not record DLco. Individuals were genotyped using the Affymetrix 6.0 SNP array.

The UK study^9^ had four centres that collected longitudinal data, namely the Brompton, PROFILE (Prospective Study of Fibrosis in the Lung Endpoints), Edinburgh and TLF (Trent Lung Function) centres. Individuals were genotyped using the Affymetrix UK BiLEVE array.

Six centres in the UUS study^5^ collected longitudinal data, namely ACE (AntiCoagulant Effectiveness in Idiopathic Pulmonary Fibrosis), Brompton, Chicago, Nottingham, PANTHER (Prednisone, Azathioprine, and N-Acetylcysteine for Pulmonary Fibrosis), and UCD (University of California, Davis) centres. The ACE and PANTHER trials did not record DLco. Individuals were genotyped using the Affymetrix UK Biobank array.

For the follow-up stage, the CleanUP-IPF (Study of Clinical Efficacy of Antimicrobial Therapy Strategy Using Pragmatic Design in Idiopathic Pulmonary Fibrosis) study^10^ was used. As CleanUP-IPF individuals were followed-up for a shorter time period than in the other three studies (up to three measures of FVC and DLco over two years), the CleanUP-IPF study was selected for the follow-up analyses and was not included in the discovery GWAS. Individuals were genotyped using the Affymetrix UK Biobank array (**Supplement**).

All studies were imputed using the Haplotype Reference Consortium^11^, had appropriate ethics approval and diagnosed cases using ATS/ERS guidelines^12,13^.

### Quality control

For quality control, we excluded individuals who failed Affymetrix genotyping quality measures, were not IPF cases, where their genetic sex did not match their recorded sex at birth, were heterozygosity outliers, non-European ancestry based on genetic principal components, duplicates and up to 2^nd^ degree related individuals. We only included individuals with at least two longitudinal measures. Where duplicates or related individuals were identified, the individual with the more complete phenotype data was kept (where this was the same, the individual from the smaller study was kept).

In our analysis we took enrolment to the study as a proxy for time of diagnosis. Some historical measures of FVC and DLco were available, however we excluded measures that were more than a year before enrolment. As most centres only recorded longitudinal measures for 3-5 years, to reduce biases in fitting longitudinal models with sparse data at later time points, we only included FVC and DLco measures taken within 3 years of enrolment. We only included variants with a minor allele frequency (MAF) greater than 1%.

### Genome-wide analysis

We performed genome-wide analyses using a longitudinal linear mixed model with random slope and intercept with a (*Time*×*SNP*) interaction term and adjusting for age, sex, the first 10 genetic principal components (to account for population stratification) and study centre, as follows:

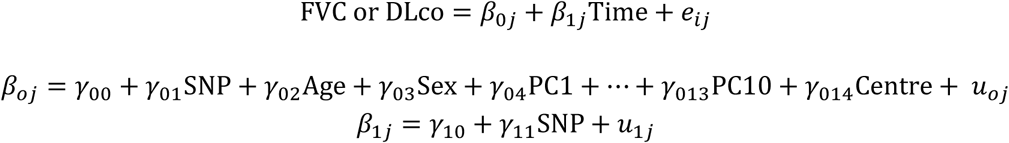

Where *e* and *u* are normally distributed random variables for level 1 and level 2 respectively. *SNP* was the genetic dosage for that variant (i.e. equal to 0 for individuals with two copies of the reference allele, 1 for heterozygotes and 2 for individuals with two copies of the effect allele), *Time* was the time in years after enrolment that the measure was taken, *Sex* was coded as 1 for males and 0 for females, *PC1* to *PC10* were the first 10 genetic principal components (included to adjust for population stratification) and *Centre* was a categorical variable for each study centre. *Age* was treated as a time varying covariate and centred on 72 (the median age of individuals at baseline). As we are interested in whether the genetic variant effects the rate of change of FVC or DLco, our effect estimate of interest is γ_11_.

The genome-wide discovery analysis was performed in each of the three studies (US, UK and UUS) separately and results were meta-analysed across studies using a fixed effect inverse variance weighted meta-analysis using the METAL software^14^. Genomic-control was applied to the meta-analysis results where λ>1. Only variants that were included in at least two of the studies were included in the meta-analysis.

Independent variants were followed-up in the CleanUP-IPF study if they reached a suggestive significance threshold in the discovery meta-analysis (p<5×10^−6^), had consistent direction of effects and reached nominal significance (p<0.05) in each of the contributing GWAS studies. Conditional analyses were performed using GCTA-COJO to identify if there were multiple independent association signals at each association locus (**Supplement**).

Variants were defined as significantly associated with FVC or DLco if they were genome-wide significant when meta-analysing across all studies (p<5×10^−8^), had consistent direction of effects and reached nominal significance (p<0.05) in each of the contributing studies.

### Gene prioritisation and characterisation of association signals

Credible sets were calculated for each associated risk signal to generate a set of variants that were 95% certain to contain the true causal variant (under the assumption there is only one causal variant and that we have measured it, **Supplement**).

To identify putative causal genes from association signals we performed the following analyses to prioritise genes of interest:

1. **Nearest gene:** The nearest gene is often found to be the gene of interest an association signal^15^, we therefore included nearest gene as part of our gene prioritisation analyses.
2. **Annotation:** To determine the functional annotation of the variants in the credible sets we used VEP^16^.
3. **Gene expression:** To determine if the association signals were associated with gene expression, we investigated whether the variants in the credible sets were associated with gene expression using publicly available eQTL resources; eQTLgen^17^ (whole blood samples from up to 31,684 individuals) and GTEx^18^ v7 (49 tissues including lung from between 73 to 706 individuals). Colocalisation analyses were performed to determine if the same variant was likely to be driving the association with FVC or DLco decline and gene expression (**Supplement**).
4. **Physical DNA interactions:** To identify genes that lie in regions of the DNA that physically interact with the region of DNA showing an association with either FVC or DLco decline, we utilised the HUGIN Hi-C database^19^ (**Supplement**).
5. **Identification of relevant Mendelian diseases:** As we hypothesise that genes associated with relevant phenotypes are more likely to be the gene of interest, we used online resources (Orphanet^20^ and OMIM [Online Mendelian Inheritance in Man]^21^) to identify nearby genes associated with relevant Mendelian diseases (**Supplement**).
6. **Rare variant associated diseases:** To identify genes associated with relevant respiratory or fibrotic phenotypes through rare variant changes or accumulation of rare variants, we investigated nearby genes using the AstraZeneca PheWAS Portal^22^ (**Supplement**).
7. **Mouse knockout models:** We investigated genes using the International Mouse Phenotyping Consortium Web Portal^23^ to identify nearby genes that exhibit relevant phenotypes when knocked-out in mice (**Supplement**).

Variants in the credible set were investigated for whether they had been previously reported as associated with any other trait in previous GWAS (**Supplement**). We also investigated whether these variants showed an association with changes in lung function in the general population (**Supplement**) and investigated the effect these variants had on IPF susceptibility in a GWAS of 4,125 IPF cases vs 20,464 controls^7^. Variants previously reported as associated with IPF susceptibility were investigated for their effect on FVC and DLco decline.

Finally, the combined effect of multiple variants in a gene and enrichment in biological pathways was tested using Vegas2^24^ (**Supplement**).

### Sensitivity analyses

To investigate variants associated with either FVC or DLco decline further, we performed the following sensitivity analyses:

1. **Short-term progression:** We repeated the longitudinal mixed model analysis only including data within 1-year of diagnosis.
2. **Clinical utility:** To assess the clinical utility of associated variants we calculated the 1-year trend of FVC for each individual (in terms of %change) and classified individuals as “progressive” if they had a 1 year decline in FVC≥10% or died within the first year. We then fit a logistic regression model to test the association between the genetic variant and this binary trait (**Supplement**).
3. **Non-linear effects:** We investigated non-linear effects for time by allowing for polynomial time and interaction effects (**Supplement**).
4. **Baseline lung function:** We tested whether the variant was associated with the first measure of FVC or DLco using a linear regression model (**Supplement**).
5. **Effect in general population:** Using 32,013 unrelated European individuals in UK Biobank, we tested whether the variant was associated with a decline of FVC, forced expiratory volume in 1 second (FEV_1_) or 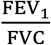 (**Supplement**).
6. **Effect of drop-out:** We tested the association of the variant with survival times using a Cox Proportional Hazards model (**Supplement**) and then performed a joint model combining the longitudinal linear mixed model and the Cox proportional hazards model (**Supplement**).
7. **Age at baseline:** To investigate whether the variant was associated with age at baseline using a linear regression model (**Supplement**).
8. **Treatment response:** Longitudinal analyses were performed allowing for an interaction between treatment, genetic variant and time (**Supplement**). Analyses were performed using the CleanUp-IPF study and the effect of associated genetic variants on nintedanib, pirfenidone and anti-microbial therapy response were investigated.

In all circumstances, sensitivity analyses were performed in the UK, UUS, US and CleanUP-IPF studies separately and then meta-analysed across studies.

## Results

There were 1,329 individuals passing quality control measures for the FVC longitudinal analysis and 975 individuals passing quality control measures for inclusion in the DLco longitudinal analysis (**Supplementary Figure 1**). There were 711 individuals included in both analyses, 337 individuals included in only the FVC analysis and 18 individuals included only in the DLco analysis. In total, there were 5,216 measures of FVC and 3,361 measures of DLco (**Table 1, Supplementary Figures 2 and 3**).

**Table 1:** Demographics of individuals included in FVC and DLco analyses.

| i) FVC | Discovery GWAS |  |  | Follow-up | Total |
| --- | --- | --- | --- | --- | --- |
|  | US | UK | UUS | CleanUP-IPF |  |
| Sample size | 142 | 314 | 592 | 281 | **1,329** |
| Study centres | 3 | 4 | 6 | 1 | **14** |
| Males | 108<br>(76.1%) | 224<br>(71.3%) | 449<br>(75.8%) | 224<br>(79.7%) | **1,005**<br>**(75.6%)** |
| Age (mean at baseline) | 65.7 | 71.7 | 68.8 | 71.3 | **69.7** |
| Total number of visits | 604 | 1,440 | 2,516 | 656 | **5,216** |
| Mean number of visits per person | 4.3 | 4.6 | 4.3 | 2.3 | **3.9** |
| Maximum number of visits | 13 | 12 | 14 | 3 | **14** |
| Number of deaths | 27<br>(19.0%) | 110<br>(35.0%) | 136<br>(23.0%) | 11<br>(3.9%) | **284**<br>**(21.4%)** |

| ii) DLco | Discovery GWAS |  |  | Follow-up | Total |
| --- | --- | --- | --- | --- | --- |
|  | US | UK | UUS | CleanUP-IPF |  |
| Sample size | 75 | 293 | 361 | 246 | **975** |
| Study centres | 2 | 4 | 4 | 1 | **11** |
| Males | 60<br>(80%) | 214<br>(73.0%) | 273<br>(75.6%) | 192<br>(78.1%) | **739**<br>**(75.8%)** |
| Age (mean at baseline) | 67.5 | 71.5 | 69.2 | 71.1 | **70.2** |
| Total number of visits | 251 | 1,146 | 1,398 | 566 | **3,361** |
| Mean number of visits per person | 3.3 | 3.9 | 3.9 | 2.3 | **3.4** |
| Maximum number of visits | 7 | 11 | 13 | 3 | **13** |
| Number of deaths | 17<br>(22.7%) | 94<br>(32.1%) | 112<br>(31.0%) | 9<br>(3.7%) | **232**<br>**(23.8%)** |

There were 7,611,174 variants included in the FVC longitudinal analysis and 7,536,843 variants in the DLco longitudinal analysis (**Figure 1** and **Supplementary Figure 4**). There were 24 independent variants in the FVC analysis and 30 independent variants in the DLco analysis reaching suggestive significance that were investigated further in the CleanUP-IPF dataset (**Supplementary Table 1**). No variants reached suggestive significance in both the FVC and DLco analyses (**Supplementary Figure 5**).

**Figure 1:**
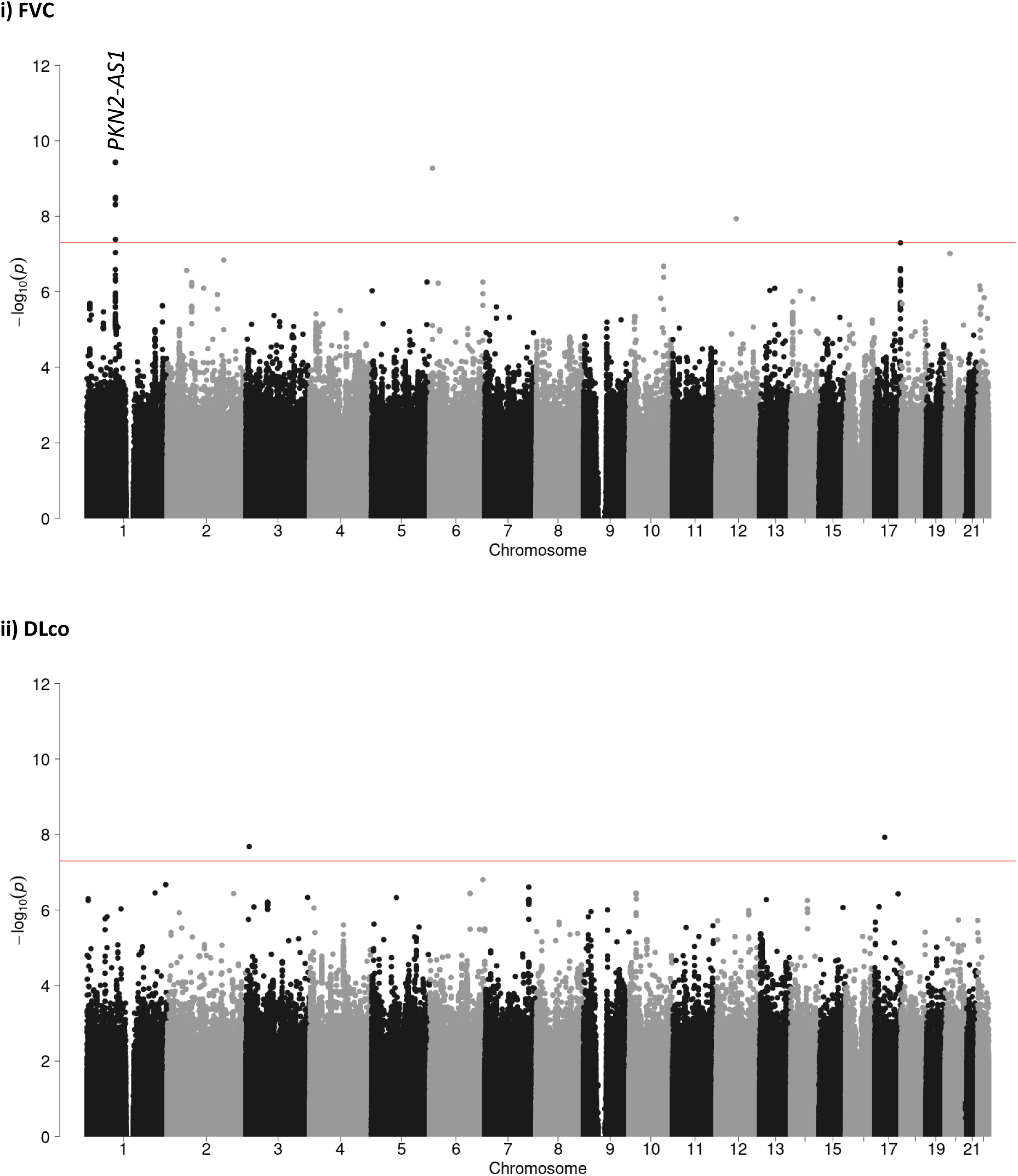
Manhattan plots for longitudinal FVC and DLco genome-wide meta-analyses. Each point represents a variant with chromosomal position on the x axis and –log(p value) on the y axis. The red horizontal line shows the genome-wide significance threshold of p=5×10^−8^.

One variant, rs115982800, met genome-wide significance (p=3.68×10^−8^) in the discovery GWAS meta-analysis with consistent direction of effects and nominal significance (p<0.05) in all discovery cohorts (**Figure 2**). This variant was also significant in CleanUP-IPF (p=0.007) with a consistent effect. Following meta-analysis of the discovery GWAS and CleanUP-IPF, this variant was associated with an annual FVC decline of 140ml/year per copy of the risk allele A (95% CI [−180, −100], p=9.14×10^−12^, **Supplementary Figure 6**). The credible set for this association signal contained seven genetic variants with three highly correlated variants accounting for 25.7% of the posterior probability each (**Supplementary Table 2)**. All of these variants were located on chromosome 1 upstream of the Protein Kinase N2 gene (*PKN2*) in introns of the antisense-RNA *PKN2-AS1* (**Figure 3**). One of the three variants in the credible set with high posterior probability (rs115590681) was located in an open chromatin region.

**Figure 2:**
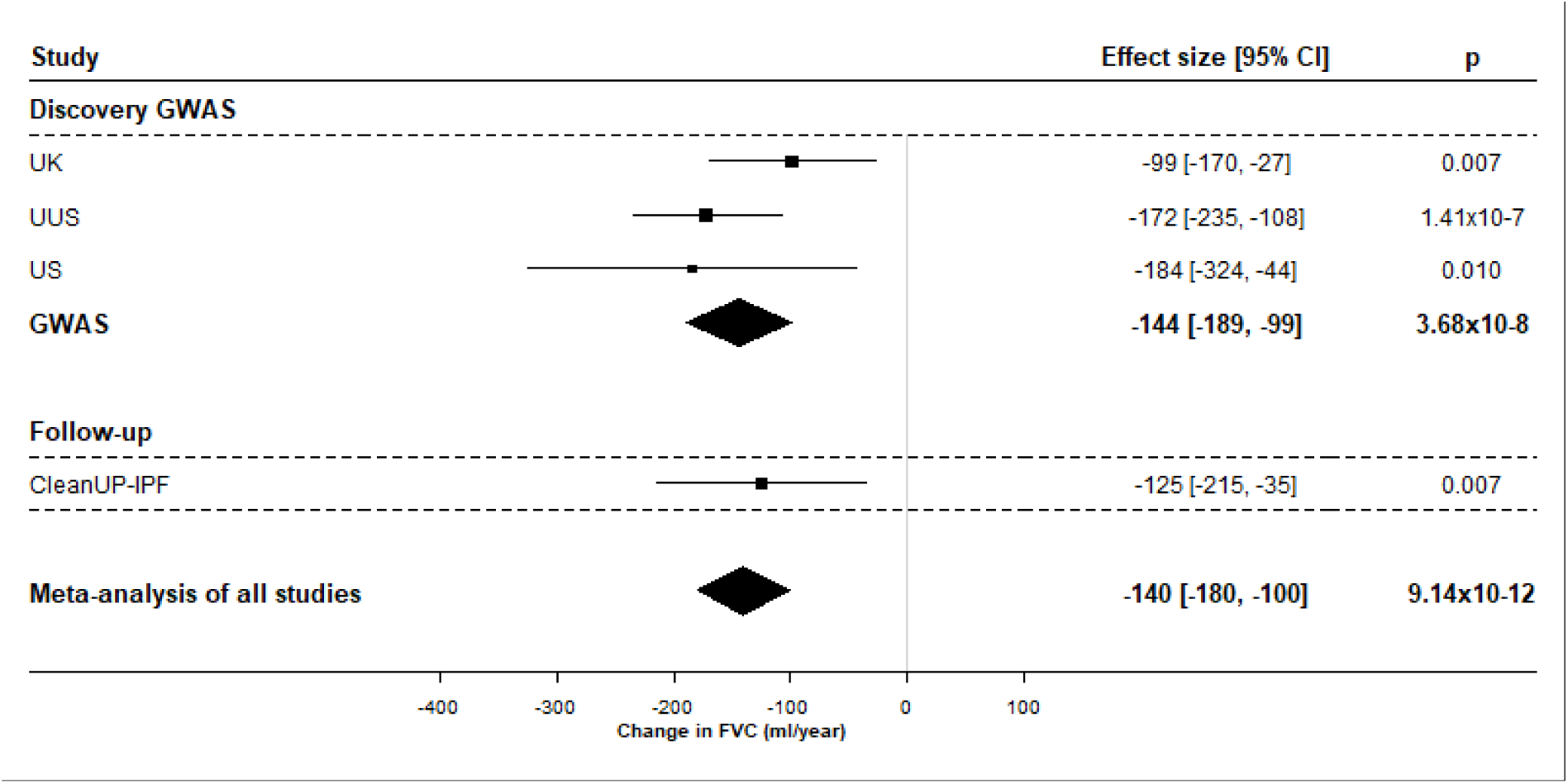
Forest plot for rs115982800.

**Figure 3:**
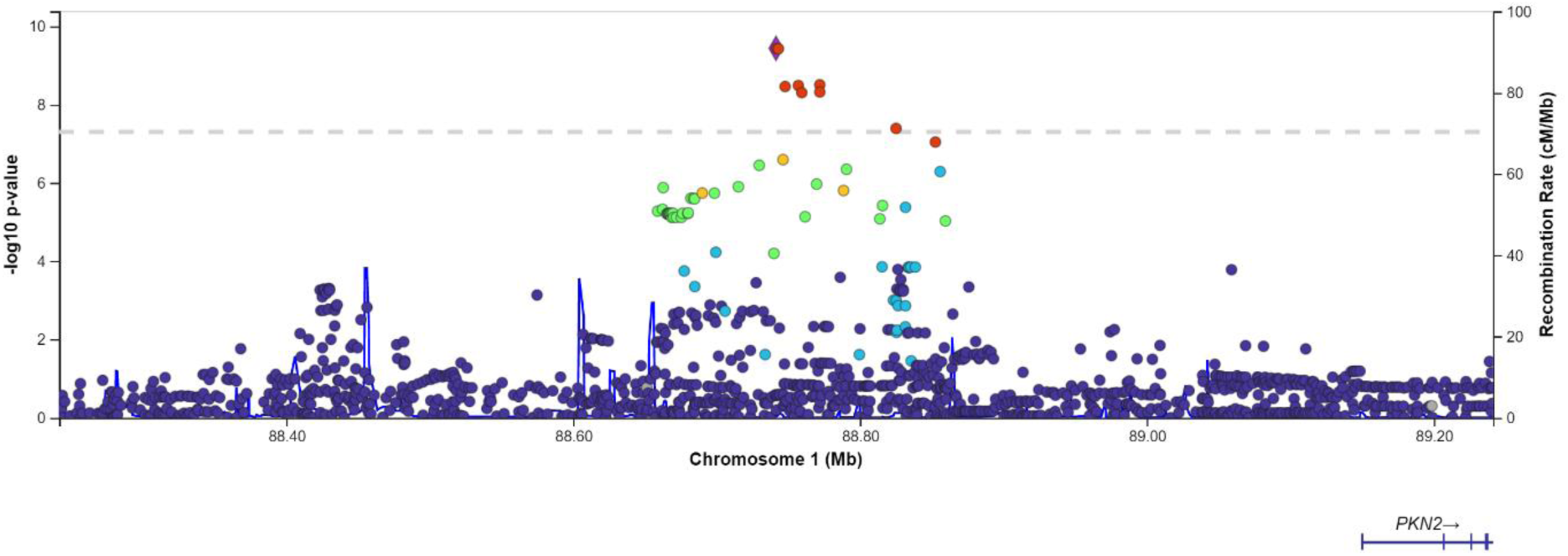
Region plot for the locus significantly associated with FVC decline. Each point represents a variant with chromosomal position on the x axis and –log(p value) on the y axis. The dashed horizontal line shows the genome-wide significance threshold of p=5×10^−8^. Variants are coloured by linkage disequilibrium with the sentinel variant (rs115982800) which is shown in blue. Variants in high linkage disequilibrium (LD) with the sentinel variant (r^2^≥0.8) are shown in red, variants with 0.8>r^2^≥0.6 are shown in orange, variants with 0.6>r^2^≥0.4 are shown in green, variants with 0.4>r^2^≥0.2 are shown in light blue and variants in weak LD with the sentinel variant (r^2^<0.2) are shown in dark blue. Light blue lines show the recombination rate and gene positions are shown below the x axis. The plot was created using LocusZoom^25^.

Sensitivity analyses showed that the association with rs115982800 remained when restricting measurements to one year after diagnosis and showed an association with clinically defined “progressive IPF” (**Supplementary Figure 7**). There was evidence of non-linear effects in the UUS study only (**Supplementary Figure 7**). The variant rs115982800 showed no association with survival times, however incorporating survival as part of a joint model did reduce the estimated effect of rs115982800 on longitudinal FVC (**Supplementary Figures 7 and 8**). The variant showed no association with baseline FVC, baseline DLco or with age at baseline (**Supplementary Figure 7**). There was no evidence of a treatment interaction between rs115982800 and response to nintedanib, pirfenidone or anti-microbial therapy (**Supplementary Table 3**), however the rs115982800 variant does show an association with Dothiepin medication in UK Biobank. The variant shows no association with IPF risk or with lung function decline in UK Biobank (**Supplementary Table 4**).

Given that the genome-wide significant association signal may be involved in long-distance gene regulation through an open chromatin, we performed gene-prioritisation on all genes within 3Mb of the sentinel variant. *PKN2* was the nearest gene and the location of the association signal in an antisense-RNA gene for *PKN2* implicated *PKN2* as a gene of interest. The fragment of DNA that contains the longitudinal FVC association signal appears to physically interact with many other parts of the DNA across multiple tissues and cell-types (**Supplementary Figure 9**). Both *PKN2* and *GBP5* are included on a diagnostic panel of 44 autoinflammatory diseases (which includes interstitial lung disease autoinflammatory disorders) but are not directly linked to a specific ILD (**Supplementary Table 5**). Rare variants in both *PKN2* (% predicted FEV_1_) and *LMO4* (FEV_1_ and FVC) show a suggestive association (p<5×10^−6^) with lung function and rare variants in *HFM1* show an association with IGF-1 levels (which is known to promote pulmonary fibrosis) (**Supplementary Table 5**). The FVC association signal did not colocalise with expression of any genes and no genes showed a relevant phenotype when knocked out in mice models (**Supplementary Figure 10 and Supplementary Table 5**). Collectively, *PKN2* had the strongest evidence of being the gene that this association signal acts through (**Supplementary Figure 11**).

Variants previously reported as associated with IPF risk did not show an association with FVC or DLco decline (**Supplementary Table 6**).

When utilising the genome-wide summary statistics, no genes or pathways were found to be significantly enriched in genetic associations with FVC or DLco decline after Bonferroni corrections for multiple testing (**Supplementary Tables 7 and 8** and **Supplementary Figure 12**). The gene *TMEM105* showed a borderline significant association with FVC decline in the gene-based analysis, however this gene includes variants in the FVC decline association signal on chromosome 17 that was not supported in the analysis in CleanUP-IPF (**Supplementary Table 1**). “GO:0051302_regulation_of_cell_division” was the only pathway term with empirical p<10^−4^ in the FVC or DLco analysis (genes in this pathway were enriched with DLco decline, p=8.80×10^−5^). This pathway contained 271 genes, many known to be associated with IPF risk including growth factor regulating genes (especially for TGFβ signalling), spindle assembly genes and WNT signalling (**Supplementary Table 8**).

## Discussion

By performing the first genome-wide association study of decline in lung health in individuals diagnosed with IPF, we have identified a genetic variant associated with declining lung capacity in individuals with IPF which lies within an antisense RNA for *PKN2*.

*PKN2* (Protein Kinase N2), which had the strongest evidence of being the causal gene for the association signal, is a Rho/RAC effector protein known to regulate cell cycle progression, actin cytoskeleton assembly, cell migration, cell adhesion, tumour cell invasion and transcription activation signalling processes^26^. RhoA is associated with IPF susceptibility through regulating TGFβ signalling and other RhoGEFs for RhoA have been implicated in GWAS of IPF risk (for example AKAP13)^9^. Protein analyses have shown PKN2 is linked to fibrotic processes in chronic atrial fibrillation^27^. IPF is a chronic lung disease characterised by fibroblast proliferation, activation and differentiation into myofibroblasts. Recent studies have shown mouse embryonic fibroblasts to depend on PKN2 for proliferation, growth and motility^28^, and cancer associated fibroblasts to depend on PKN2 for activation, differentiation and motility^29^. In addition, PKN2 has been suggested to play critical roles in actin stress fibre formation in NIH3T3 cells^30^. PKN2 inhibitors are currently in development for cancer therapy^31^ and the PKN2 inhibitor Fostamatinib has been suggested as a drug repurposing candidate for the treatment of acute respiratory distress syndrome in severe COVID-19 patients^32^.

The variant we identified as associated with FVC decline also shows an association with Dothiepin medication. Dothiepin is an anti-depressant that has previous been reported as a potential risk factor for IPF^33^.

There are a number of strengths to this study. Firstly, the identified association signal shows a consistent association and effect size estimate across four independent studies, suggesting this variant is robustly associated with FVC decline in individuals with IPF. Secondly, sensitivity analyses showed that the signal remained when restricting FVC measurements to the first year after diagnosis and the variant also showed an association with clinically defined “progressive IPF”. Thirdly, by applying a mixed model, we were able to maximise the power of the analysis by incorporating over 5,000 measures of FVC and 3,000 measures of DLco. We assumed a linear mixed effects model as we were not aiming to model lung function trajectories in IPF individuals, rather we were interested in genetic variants associated with declining FVC and DLco. Although it is unlikely that FVC or DLco trajectories follow a linear trend, assuming linearity simplifies the model (increasing power and aiding model convergence) while still being able to distinguish between individuals with a generally declining trend against those with a relatively stable disease. Sensitivity analyses did show a nominal association with non-linear effects though the effect size was small and a significant non-linear effect was only observed in one study.

There are also limitations to the analysis. Firstly, our model assumes non-informative dropout which is unlikely to be true, especially for DLco. Sensitivity analyses showed that although rs115982800 is not associated with survival times, incorporating censoring due to death through a joint model does remove the association with longitudinal FVC meaning censoring due to death may have some effect on the analysis. The joint model coefficients are estimated together and it is possible the effect of the variant is being distributed across the longitudinal and time-to-event parts of the model. This is especially true given the complexity of the joint model and if rs115982800 explains little of the variance. Furthermore, rs115982800 showed consistent effect size estimates across all four studies with varying levels of censoring, including the CleanUP-IPF study where there was very little censoring, suggesting this association is not purely caused by informative dropout. Secondly, the credible set included a variant lying in an open chromatin region and multiple regions were significant in the Hi-C analyses. This suggests the signal could be involved in regulating a large number of genes in a tissue or cell-specific manner. Thirdly, to reduce confounding we only included European individuals meaning we may miss genetic variants associated with progressive IPF. The rs115982800_A allele is low frequency in most populations and rare in African or Asian populations^34^. Finally, although we have maximised the available power of the analysis by incorporating multiple measurements, larger studies are needed to detect variants with smaller effect sizes or for rarer variants associated with lung function decline in individuals with IPF. The smaller sample size analysis means there was lower statistical power in the DLco analysis, which may explain why no association signals were identified.

In summary, by utilising thousands of lung health measures in individuals we have shown a genetic association with worsening lung capacity in IPF. These results highlight the role of protein kinase N2 in disease progression and may aid the development of new and desperately needed treatments.

## Data Availability

Full summary statistics for the FVC and DLco genome-wide meta-analyses can be accessed from https://github.com/genomicsITER/PFgenetics.

## Ethics statement

This research was conducted using previously published work with appropriate ethics approval. The PROFILE study (which provided samples for the UK and UUS studies) had institutional ethics approval at the University of Nottingham (NCT01134822 – ethics reference 10/H0402/2) and Royal Brompton and Harefield NHS Foundation Trust (NCT01110694 – ethics reference 10/H0720/12). UK samples were recruited across multiple sites with individual ethics approval (University of Edinburgh Research Ethics Committee [The Edinburgh Lung Fibrosis Molecular Endotyping (ELFMEN) Study NCT04016181] 17/ES/0075, and Nottingham Research Ethics Committee 09/H0403/59). For individuals recruited at the University of Chicago, consenting patients with IPF who were prospectively enrolled in the institutional review board-approved ILD registry (IRB#14163A) were included. Individuals recruited at the University of Pittsburgh Medical Centre had ethics approval from the University of Pittsburgh Human Research Protection Office (reference STUDY20030223: Genetic Polymorphisms in IPF). This study also included individuals from clinical trials with ethics approval (ACE [NCT00957242], PANTHER [NCT00650091], COMET [NCT01071707] and CleanUP-IPF [NCT02759120] studies).

## Author contributions

RJA, JMO, DAJ, OCL, BGG and CAM performed the analyses. JMO, SFM, JJ, JSK, WAF, EO, RBH, VN, RB, GS, MDT, NH, MKBW, NK, YZ, FJM, ALL, AA, MES, TMM, PLM, CF, IN, RGJ and LVW recruited individuals, genotyped samples and collected longitudinal phenotypic data. RJA, DAJ and LVW developed the analysis plan and CF, IN, RGJ and LVW supervised the study. RJA, JMO, CF, IN, RGJ and LVW wrote the first draft of the manuscript and all authors contributed and edited the final version.

