## Supplement for "Genome-wide analysis of longitudinal lung function and gas transfer in individuals with idiopathic pulmonary fibrosis"

Supplementary Figure 11: Summary of gene prioritisation analyses. 42
Supplementary Figure 12: Manhattan plots for gene-based analyses 43
i) FVC 43
ii) DLco 43
Supplementary references 44

### Supplementary methods

#### Genotyping and quality control of the CleanUP-IPF study

A total of 513 IPF cases from the CleanUP-IPF study were genotyped alongside 51 IPF cases recruited at the University of California, Davis (UCD) and 12 Affymetrix control samples. Genotyping was performed using the Affymetrix UK Biobank array and genotype calling was performed by Affymetrix using the Axiom Analysis Suite.

There were 836,727 variants genotyped. Variants were excluded pre-imputation for the following reasons; 10,799 were removed for there being a better probe set, 16,757 failed clustering QC and 562 had a variant call rate < 95%.

Four individuals (one failed dish QC and a further three had a call rate < 97% in the first step genotype calling) were removed by Affymetrix. After the second step calling no individuals were removed for having a call rate < 95%.

Genetic principal components were calculated using PLINK v1.9 on a pruned set of variants passing the following criteria: genotyping call rate > 95%, autosomal, minor allele frequency > 1%, were in Hardy-Weinberg equilibrium ($p > 10^{-6}$), not in regions of high linkage disequilibrium (LD) and included in the HapMap samples. This was then merged with unrelated HapMap samples and principal components were calculated. K means clustering was then performed, changing the value of k, until a cluster was generated that clearly identified a cluster of European HapMap samples. All IPF cases within the European cluster were defined as Europeans. A value of k=7 was found to be the best value for clustering. Only European individuals were included in the longitudinal analysis.

Ancestry adjusted heterozygosity rates were calculated by regressing the first four principal components (and interactions) on the unadjusted heterozygosity rate. There were no individuals excluded for having a high heterozygosity rate (> 5 standard deviations above the mean).

We identified duplicates and related individuals within the CleanUP-IPFsamples, and between the other studies included in the longitudinal GWAS using KING[1]. Duplicates were defined as pairs of individuals with kinship > 0.3540, first-degree relatives with kinship [0.1770, 0.3540], second-degree relatives with kinship [0.0884, 0.1770] and third-degree relatives with kinship [0.0442, 0.0884]. This analysis identified one individual as a third-degree relative of a large number of other unrelated individuals. This individual was removed (although this individual did not meet the exclusion criteria based on heterozygosity, they did show high levels of heterozygosity before adjusting for ancestry). For the longitudinal GWAS, where duplicates or related individuals (up to second-degree) were identified (across any study), the individual with more longitudinal measures was kept in the analysis and the other individual was removed. If the same number of visits was available, the individual from the smaller study was kept.

Genetic sex was inferred using PLINK and by Affymetrix. Where this differed with the recorded sex, the actual sex of the individual was checked and where sex mismatches remained, these individuals were removed as it suggests there may be sample mix-up. Seven individuals were removed for being sex mismatches.

The individuals passing quality control were imputed to the Haplotype Reference Consortium using the Michigan Imputation server[2,3]. Cases were imputed alongside 2,455 unrelated European controls who had been selected from UK Biobank such that there were five controls per case and such that cases and controls followed similar sex and smoking distributions. Controls were not included in this study of longitudinal FVC or DLco.

### Conditional analyses

Conditional analyses were performed to identify independent association signals using the meta-analysis summary statistics using GCTA-COJO[4]. Independent signals were defined as signals meeting the significance threshold as defined in the main analysis (i.e. meta-analysis $p < 5 \times 10^{-8}$, consistent direction of effects and $p < 0.05$ in each of the studies contributing to the meta-analysis), after conditioning on the most associated variant in the region.

### Gene prioritisation and characterisation of association signals

#### Credible sets

Credible sets were calculated for each novel signal to produce a set of variants likely to contain the causal variant at 95% confidence. This method assumes there is a single causal variant and that variant had been measured. Posterior probabilities of the variant being causal were calculated for all variants within 1Mb of the sentinel variant and in at least weak linkage disequilibrium with the sentinel variant ($r^2 > 0.1$). Posterior probabilities were calculated from approximate Bayes factors (ABFs) using the formula proposed by Wakefield[5]:

$$\mathrm{ABF} = \frac{1}{\sqrt{1 - \frac{W}{V+W}}} \exp\left(-\frac{Z^2}{2}\frac{W}{V+W}\right)$$

where $W$ is the Wakefield prior (which we set to 0.04 as this is equivalent to a 95% belief that departure from the null model for the relative risk is less than 1.5), $Z$ is the Z statistic for the variant and $V$ is the variance of the effect size.

The approximate posterior probability was set to equal the ABF for that variant divided by the sum of ABFs for all variants in the signal. Variants were added to the credible set until the sum of the posterior probabilities was greater than or equal to 0.95.

#### Colocalisation between FVC and DLco signals with gene expression

Variants in the 95% credible sets were tested for their association with gene expression in three publicly available eQTL (expression quantitative trait loci) databases; namely eQTLGen, GTEx and the lung eQTL database. eQTLGen[6] contains blood eQTL data from 31,684 individuals meta-analysed across 37 studies. GTEx (Genotype-Tissue Expression project)[7] v8 has samples from between 73 and 706 individuals for 49 tissues, including whole lung. The lung eQTL database consisted of lung samples from 1,111 individuals from Universities of British Columbia, Laval and Groningen[8-10]. A false discovery rate threshold of 5% was used for the eQTLGen and GTEx studies and 10% for the lung eQTL study.

Where IPF susceptibility variants were found to be associated with expression levels of a gene, we tested whether the same variant was likely to be causal both for differences in gene expression and

IPF susceptibility. Colocalisation analyses were performed following the approach proposed by Giambartolomei et al[11] using the coloc package in R v3.6.1. In summary, it uses approximate Bayes factors to estimate the probability of each of the following models:

- $H_o$: There is no association in the region with either longitudinal lung function in IPF or the eQTL analysis
- $H_1$: There is an association in the region with longitudinal lung function in IPF but not with the expression of the gene
- $H_2$: There is an association in the region with the expression of the gene but not with longitudinal lung function in IPF
- $H_3$: There is an association in the region with both longitudinal lung function in IPF and the expression of the gene but these are driven by two different variants
- $H_4$: There is an association in the region with both longitudinal lung function in IPF and the expression of the gene, which is driven by the same variant.

We took colocalisation to be when the probability of $H_4$ (i.e. the same variant drives longitudinal decline in FVC or DLco and the expression of the gene) was greater than 80%.

**Physical DNA interactions**

To investigate whether the DNA fragment that contains the association with FVC or DLco decline physically interacts with other genomic regions we used the Hi-C Unifying Genomic Integrator (HUGIn)[12]. Physical interactions within 3Mb of the fragment region containing the sentinel association variant were investigated in seven human cell lines (lymphoblastoid cell, embryonic stem cell, fetal lung fibroblast cell, Mesendoderm cell, mesenchymal stem cell, neural progenitor cell and trophoblast-like cell) and 14 human primary tissues (adrenal, aorta, bladder, dorsolateral prefrontal cortex, hippocampus, lung, liver, left ventricle, right ventricle, ovary, pancreas, psoas, small bowel and spleen). Physical interactions were defined as significant if they met the HUGIn defined Bonferroni corrected significance threshold. Note, the fragments of DNA used were quite large (40kB wide) and it is not possible to determine which part of the fragments the physical interactions are occurring. A fragment can contain several genes and a gene may be split across several fragments.

**Identification of relevant Mendelian diseases**

To investigate if nearby genes have been reported as associated with relevant phenotypes, we searched the genes using Online Mendelian Inheritance in Man (OMIM)[13]. OMIM is a daily updated compendium on Mendelian disorders covering 16,000 genes.

**Rare variant associated diseases**

To test whether rare variants in nearby genes have been reported as associated with relevant diseases, we investigated the genes using Orphanet[14]. Rare variants in nearby genes were also investigated using the AstraZeneca PheWAS Portal[15]. Both single variant and gene-based tests were investigated for their association with up to 18,780 traits in UK Biobank, and deemed to show an association with a trait if it met the suggestive significance threshold of $p < 10^{-6}$.

**Mouse knockout models**

To investigate whether knocking out any nearby genes results in relevant phenotypes, we investigated the genes using the International Mouse Phenotype Consortium[16] publicly available data.

### Phenome-wide association study

To investigate whether the variants associated with FVC or DLco decline in IPF patients are associated with other traits.

We used the Open Targets Genetics portal to investigate whether any of the variants in the credible set had been previously reported as associated with any other traits (at genome-wide significance level $p < 5 \times 10^{-8}$) in GWAS Catalog or in automated analyses of the UK Biobank and FinnGen general population biobanks.

To investigate whether the variants were associated with IPF specific traits, we tested the variants association with IPF susceptibility in a GWAS of 4,125 IPF cases vs 20,464 controls[17].

### Gene-based and pathway analyses

Gene-based tests were performed using Vegas2. This method combines the summary statistics from the genome-wide analysis for variants located within a gene. Variants were pruned using the UUS study as an LD reference panel (the stringency of the pruning varies depending on the number of variants in the gene). A randomly simulated vector of test statistics, with mean 0, is generated based on the distribution of observed test statistics for variants in that gene. The observed test statistics are then tested against the simulated test statistics. These simulations are then repeated and the gene-based p value is calculated based on the proportion of times the observed test statistics are statistically different from the randomly simulated test statistics.

A similar approach is used for testing for enrichment in biological pathways using the gene-based test statistics. Genes were assigned to pathways using the Biosystems annotation file available on the Vegas2 website (https://vegas2.qimrberghofer.edu.au/, accessed 24th March 2021), and the p value distributions of the genes in the pathway were tested against simulated test statistics.

Genes were defined as significantly associated if the p value met a Bonferroni corrected p threshold. Due to many pathways containing the same genes, pathways were defined as significantly enriched if the empirical p value $< 10^{-5}$.

### Sensitivity analyses for rs115982800

The only variant reaching our significance threshold in the GWAS study was rs115982800 where the minor allele showed an association with decreasing FVC. We performed sensitivity analyses to investigate this association further and to test assumptions of the model. In all equations below, the variable *SNP* is equal to 0 if the individual was homozygous for the major allele (GG) for rs115982800, 1 if the individual was heterozygous (AG) and 2 if the individual had two copies of the minor allele (AA).

#### Short term progression

As linear models may be more appropriate for modelling short term progression in IPF, we repeated the longitudinal analysis using the same linear mixed model, using only FVC measures taken within 1 year of diagnosis.

#### Clinical utility

Progressive IPF is commonly defined as a decrease in FVC by 10% in the first year or death within 1 year. FVC values were recalculated as % values of the first FVC measurement and a line of best fit for

FVC was fit for each individual. If this slope showed a ≥ 10% decline in FVC over the first year or the individual died within 1 year of diagnosis, the individual was defined as “progressive”. The following logistic regression model was used to test whether rs115982800 was associated with clinically defined progressive IPF:

$$\log\left(\frac{p}{1-p}\right) = \beta_0 + \beta_1\text{SNP} + \beta_2\text{Age} + \beta_3\text{Sex} + \beta_4\text{PC1} + \cdots + \beta_{13}\text{PC10} + \beta_{14}\text{Centre} + e$$

where p is the probability of being “progressive”.

**Non-linear effects**

The longitudinal model we fit for the genome-wide analysis assumes a linear change in FVC decline. To relax this assumption, we firstly allowed quadratic *Time* effects, i.e.:

$$\text{FVC} = \beta_{0j} + \beta_{1j}\text{Time} + \beta_{2j}\text{Time}^2 + e_{ij}$$

$$\beta_{oj} = \gamma_{00} + \gamma_{01}\text{SNP} + \gamma_{02}\text{Age} + \gamma_{03}\text{Sex} + \gamma_{04}\text{PC1} + \cdots + \gamma_{013}\text{PC10} + \gamma_{014}\text{Centre} + u_{oj}$$
$$\beta_{1j} = \gamma_{10} + \gamma_{11}\text{SNP} + u_{1j}$$
$$\beta_{2j} = \gamma_{20} + \gamma_{21}\text{SNP} + u_{2j}$$

where the interaction between *SNP* and linear *Time* ($\gamma_{11}$) and quadratic *Time* ($\gamma_{21}$) will be the effect estimates of interest. Secondly, we allowed for cubic *Time* effects:

$$\text{FVC} = \beta_{0j} + \beta_{1j}\text{Time} + \beta_{2j}\text{Time}^2 + \beta_{3j}\text{Time}^3 + e_{ij}$$

$$\beta_{oj} = \gamma_{00} + \gamma_{01}\text{SNP} + \gamma_{02}\text{Age} + \gamma_{03}\text{Sex} + \gamma_{04}\text{PC1} + \cdots + \gamma_{013}\text{PC10} + \gamma_{014}\text{Centre} + u_{oj}$$
$$\beta_{1j} = \gamma_{10} + \gamma_{11}\text{SNP} + u_{1j}$$
$$\beta_{2j} = \gamma_{20} + \gamma_{21}\text{SNP} + u_{2j}$$
$$\beta_{3j} = \gamma_{30} + \gamma_{31}\text{SNP} + u_{3j}$$

**Baseline lung function**

We investigated whether the variant was associated with the first measure of FVC or DLco using a linear regression model adjusting for age, sex, study centre and the first 10 genetic principal components, i.e.:

$$\text{FVC or DLco} = \beta_0 + \beta_1\text{SNP} + \beta_2\text{Age} + \beta_3\text{Sex} + \beta_4\text{PC1} + \cdots + \beta_{13}\text{PC10} + \beta_{14}\text{Centre} + e$$

**Effect in general population**

We investigated whether the variants associated with FVC or DLco decline in IPF individuals were associated with changes of FVC, forced expiratory volume in 1 second ($FEV_1$) or the ratio $FEV_1$/FVC in 32,013 unrelated individuals of European ancestry from UK Biobank.

Given the computational difficulties in fitting linear mixed models to large sample sizes, the longitudinal analysis was performed using a conditional two-step approach[18]. Firstly, the longitudinal model without SNP effects was fit and the random effect of time for each individual was extracted. Secondly, the SNP was tested for its association with the random time effect using a linear regression model, with adjustment for 15 genetic principal components and genotyping array.

**Effect of drop out**
Individuals with more progressive forms of the disease are more likely to die at earlier time points. This means as time goes on, the individuals left in the analysis are more likely to have a less progressive form of IPF. We therefore, tested whether drop-out of the study due to death was causing biases of the results. Individuals still alive after three years were censored.

Firstly, we performed the following Cox proportional hazards model to test whether rs115982800 was associated with transplant-free survival times after diagnosis of IPF:

$$h(t) = h_0(t)\exp(\beta_0 + \beta_1\text{SNP} + \beta_2\text{Age} + \beta_3\text{Sex} + \beta_4\text{Centre})$$

where *h(t)*, the hazard function, is the rate of death at time *t* and $h_0(t)$ is the baseline hazard function. All individuals were censored at 3 years if still in the study.

Secondly, we fit a joint model of the longitudinal and survival models to investigating whether incorporating drop-out affects the longitudinal results. For this the coefficients are estimated by maximising the joint distribution of the longitudinal and survival models.

$$\text{FVC } = \beta_{0j} + \beta_{1j}\text{Time} + e_{ij}$$

$$\beta_{oj} = \gamma_{00} + \gamma_{01}\text{SNP} + \gamma_{02}\text{Age} + \gamma_{03}\text{Sex} + \gamma_{04}\text{Centre} + \ u_{oj}$$
$$\beta_{1j} = \gamma_{10} + \gamma_{11}\text{SNP} + u_{1j}$$

$$h(t) = h_0(t)\exp(\alpha_0 + \alpha_1\text{SNP} + \alpha_2\text{Age} + \alpha_3\text{Sex} + \alpha_4\text{Centre})$$

As with the main analysis, we are interested in whether the variant affects the slope of FVC over time and $\gamma_{11}$ is the variable of interest.

**Note:** Due to the complexity of the joint model, genetic principal components were not included to help the model converge.

**Age at baseline**
We investigated whether the variant was associated with age at baseline using a linear regression model adjusting for sex, study centre and the first 10 genetic principal components, i.e.:

$$\text{Age} = \beta_0 + \beta_1\text{SNP} + \beta_2\text{Sex} + \beta_3\text{PC1} + \cdots + \beta_{12}\text{PC10} + \beta_{13}\text{Centre} + e$$

**Treatment response**
To test whether there was any difference in treatment response between individuals with FVC or DLco decline associated variants, we fit the longitudinal model allowing for an interaction with treatment status, i.e.:

$$\text{FVC or DLco} = \beta_{0j} + \beta_{1j}\text{Time} + e_{ij}$$

$$\beta_{oj} = \gamma_{00} + \gamma_{01}\text{SNP} + \gamma_{02}\text{Age} + \gamma_{03}\text{Sex} + \gamma_{04}\text{PC1} + \cdots + \gamma_{013}\text{PC10} + \gamma_{014}\text{Centre} + \gamma_{015}\text{Treatment} + \gamma_{016}(\text{SNP} \times \text{Treatment}) + \ u_{oj}$$

$$\beta_{1j} = \gamma_{10} + \gamma_{11}\text{SNP} + \gamma_{12}\text{Treatment} + \ \gamma_{13}(\text{SNP} \times \text{Treatment}) + u_{1j}$$

where *Treatment* is equal to 1 if the individual was being prescribed the treatment at baseline and 0 otherwise. In this instance, we are interested in whether the *SNP*×*Treatment* interaction changes the slope of FVC or DLco and therefore $\gamma_{13}$ is the effect estimate of interest.

Only the CleanUP-IPF study was used for this analysis as this was the only study with detailed medication records which recruited individuals since the IPF treatments nintedanib and pirfenidone were licenced for use. The treatment's investigated were nintedanib, pirfenidone and the antimicrobial therapy that individuals in the CleanUP-IPF study were randomised to.

### Supplementary Tables

**Supplementary Table 1: Discovery and follow-up results for variants reaching suggestive significance in discovery GWAS**

Suggestive significant variants were defined as those with $p<5\times10^{-6}$ in the genome-wide meta-analysis plus consistent direction of effects and $p<0.05$ in each of the studies contributing to the meta-analysis. Chr=Chromosome. EAF=Effect allele frequency. Positions are based on genetic build 37.

**i) FVC**

| Chr | Position | rsid | Ref allele | Effect allele | EAF | GWAS | | Follow-up | | Meta-analysis | |
|---|---|---|---|---|---|---|---|---|---|---|---|
| | | | | | | β [95% CI] | p | β [95% CI] | p | β [95% CI] | p |
| 1 | 88741347 | rs115982800 | G | A | 5.9% | -0.14 [-0.19, -0.10] | $3.68\times10^{-10}$ | -0.12 [-0.21, -0.04] | 0.007 | -0.14 [-0.18, -0.10] | $9.14\times10^{-12}$ |
| 1 | 96384437 | rs567748245 | A | T | 1.3% | -0.33 [-0.47, -0.18] | $8.75\times10^{-6}$ | 0.05 [-0.16, 0.27] | 0.624 | -0.21 [-0.33, -0.09] | $6.79\times10^{-4}$ |
| 2 | 154838235 | rs144955197 | C | G | 1.5% | -0.34 [-0.48, -0.20] | $1.19\times10^{-6}$ | -0.03 [-0.23, 0.17] | 0.755 | -0.24 [-0.35, -0.13] | $3.08\times10^{-5}$ |
| 2 | 155504296 | rs190903032 | C | T | 1.6% | -0.28 [-0.40, -0.16] | $2.86\times10^{-6}$ | -0.03 [-0.20, 0.13] | 0.705 | -0.20 [-0.29, -0.10] | $5.91\times10^{-5}$ |
| 2 | 174360794 | rs146936508 | A | G | 1.6% | -0.26 [-0.36, -0.16] | $1.45\times10^{-7}$ | 0.03 [-0.12, 0.18] | 0.679 | -0.17 [-0.26, -0.09] | $3.23\times10^{-5}$ |
| 3 | 104587601 | rs116289237 | A | G | 1.3% | 0.35 [0.20, 0.51] | $6.20\times10^{-6}$ | 0.06 [-0.13, 0.25] | 0.562 | 0.24 [0.12, 0.35] | $1.16\times10^{-4}$ |
| 4 | 94432657 | rs147204574 | A | G | 1.2% | -0.27 [-0.38, -0.15] | $3.13\times10^{-6}$ | 0.07 [-0.26, 0.40] | 0.676 | -0.23 [-0.34, -0.12] | $2.70\times10^{-5}$ |
| 5 | 2495673 | rs78622840 | C | T | 2.4% | -0.20 [-0.28, -0.12] | $9.36\times10^{-7}$ | -0.01 [-0.14, 0.12] | 0.903 | -0.15 [-0.22, -0.08] | $2.99\times10^{-5}$ |
| 5 | 171379897 | rs111327342 | A | G | 2.4% | 0.19 [0.10, 0.27] | $7.39\times10^{-6}$ | 0.02 [-0.12, 0.16] | 0.794 | 0.14 [0.07, 0.21] | $8.82\times10^{-5}$ |
| 5 | 171928664 | rs73802669 | C | A | 2.3% | 0.21 [0.12, 0.29] | $5.52\times10^{-7}$ | 0.04 [-0.08, 0.16] | 0.547 | 0.15 [0.09, 0.22] | $7.02\times10^{-6}$ |
| 6 | 8339921 | rs76782037 | T | C | 1.5% | -0.34 [-0.45, -0.23] | $5.31\times10^{-10}$ | 0.01 [-0.19, 0.22] | 0.895 | -0.27 [-0.36, -0.17] | $4.42\times10^{-8}$ |

| | | | | | | | | | | | |
|---|---|---|---|---|---|---|---|---|---|---|---|
| 6 | 164417620 | rs114052286 | C | T | 3.0% | 0.14 [0.08, 0.20] | $5.54 \times 10^{-7}$ | -0.07 [-0.22, 0.08] | 0.352 | 0.11 [0.06, 0.17] | $2.61 \times 10^{-5}$ |
| 7 | 35500852 | rs75679695 | A | G | 1.9% | -0.21 [-0.30, -0.12] | $5.02 \times 10^{-6}$ | -0.05 [-0.18, 0.08] | 0.480 | -0.15 [-0.23, -0.08] | $4.54 \times 10^{-5}$ |
| 7 | 76146001 | 7.76E+08 | T | C | 53.7% | 0.06 [0.04, 0.09] | $4.77 \times 10^{-6}$ | -0.01 [-0.05, 0.03] | 0.550 | 0.04 [0.02, 0.06] | 0.001 |
| 10 | 103043841 | rs143967606 | G | T | 1.9% | -0.21 [-0.31, -0.12] | $8.96 \times 10^{-6}$ | 0.13 [-0.04, 0.29] | 0.124 | -0.13 [-0.21, -0.05] | 0.002 |
| 11 | 20074608 | rs192969588 | C | A | 1.5% | 0.31 [0.17, 0.44] | $9.18 \times 10^{-6}$ | -0.02 [-0.22, 0.18] | 0.832 | 0.20 [0.09, 0.32] | $4.66 \times 10^{-4}$ |
| 13 | 66119518 | rs117498630 | G | A | 2.8% | 0.17 [0.11, 0.24] | $8.05 \times 10^{-7}$ | 0.00 [-0.19, 0.20] | 0.981 | 0.16 [0.09, 0.22] | $2.80 \times 10^{-6}$ |
| 16 | 11807880 | rs35478579 | G | A | 1.9% | -0.23 [-0.33, -0.13] | $7.52 \times 10^{-6}$ | -0.06 [-0.23, 0.12] | 0.524 | -0.19 [-0.27, -0.10] | $3.24 \times 10^{-5}$ |
| 17 | 79298094 | rs116943420 | G | A | 1.9% | -0.25 [-0.34, -0.16] | $5.03 \times 10^{-8}$ | - | - | - | - |
| 18 | 2882412 | rs76188719 | C | T | 1.5% | 0.23 [0.13, 0.32] | $2.05 \times 10^{-6}$ | 0.03 [-0.11, 0.16] | 0.689 | 0.16 [0.08, 0.24] | $4.08 \times 10^{-5}$ |
| 18 | 75152877 | rs11661110 | G | A | 5.6% | -0.13 [-0.18, -0.07] | $6.29 \times 10^{-6}$ | 0.07 [-0.04, 0.17] | 0.216 | -0.08 [-0.13, -0.03] | 0.001 |
| 20 | 14762116 | rs78380407 | C | G | 1.7% | -0.26 [-0.36, -0.17] | $9.68 \times 10^{-8}$ | 0.06 [-0.09, 0.20] | 0.431 | -0.16 [-0.24, -0.08] | $7.22 \times 10^{-5}$ |
| 22 | 24189485 | rs143420816 | G | A | 1.4% | -0.22 [-0.31, -0.13] | $6.97 \times 10^{-7}$ | 0.21 [-0.06, 0.48] | 0.123 | -0.18 [-0.27, -0.10] | $3.49 \times 10^{-5}$ |
| 22 | 37339064 | rs117246948 | C | G | 1.1% | 0.28 [0.17, 0.39] | $1.44 \times 10^{-6}$ | 0.04 [-0.11, 0.19] | 0.635 | 0.19 [0.10, 0.28] | $3.38 \times 10^{-5}$ |

**ii) DLco**

| Chr | Position | rsid | Ref allele | Effect allele | EAF | GWAS | | Follow-up | | Meta-analysis | |
|---|---|---|---|---|---|---|---|---|---|---|---|
| | | | | | | β [95% CI] | p | β [95% CI] | p | β [95% CI] | p |
| 1 | 4359223 | rs114768831 | G | A | 1.3% | 0.93 [0.57, 1.29] | $5.00\times10^{-7}$ | -0.05 [-0.46, 0.36] | 0.809 | 0.50 [0.23, 0.77] | $3.24\times10^{-4}$ |
| 1 | 62715361 | rs142240901 | C | T | 1.8% | 0.54 [0.32, 0.76] | $1.50\times10^{-6}$ | -0.19 [-0.79, 0.40] | 0.524 | 0.45 [0.24, 0.66] | $1.92\times10^{-5}$ |
| 1 | 172449194 | rs114203938 | T | C | 11.5% | 0.18 [0.10, 0.26] | $9.41\times10^{-6}$ | 0.03 [-0.13, 0.19] | 0.694 | 0.15 [0.08, 0.22] | $3.72\times10^{-5}$ |
| 1 | 211218391 | 1211218391 | T | G | 1.6% | -0.83 [-1.15, -0.51] | $3.51\times10^{-7}$ | -0.42 [-0.97, 0.13] | 0.134 | -0.73 [-1.00, -0.45] | $2.63\times10^{-7}$ |
| 2 | 44313803 | rs150570718 | A | G | 2.1% | -0.49 [-0.69, -0.28] | $2.97\times10^{-6}$ | -0.01 [-0.32, 0.30] | 0.937 | -0.34 [-0.51, -0.17] | $8.27\times10^{-5}$ |
| 2 | 168997340 | rs146202774 | A | T | 1.3% | -0.59 [-0.85, -0.33] | $8.54\times10^{-6}$ | -0.02 [-0.42, 0.38] | 0.921 | -0.42 [-0.64, -0.21] | $1.40\times10^{-4}$ |
| 3 | 10296133 | rs2241309 | G | A | 1.3% | -0.71 [-0.96, -0.46] | $2.08\times10^{-8}$ | 0.11 [-0.34, 0.55] | 0.643 | -0.52 [-0.73, -0.30] | $3.22\times10^{-6}$ |
| 3 | 67521429 | rs73104351 | A | T | 2.4% | 0.48 [0.29, 0.67] | $6.22\times10^{-7}$ | -0.03 [-0.39, 0.32] | 0.854 | 0.37 [0.20, 0.54] | $1.43\times10^{-5}$ |
| 3 | 191456988 | rs139607041 | C | T | 1.4% | -0.64 [-0.89, -0.39] | $4.65\times10^{-7}$ | -0.40 [-1.17, 0.37] | 0.304 | -0.62 [-0.86, -0.38] | $2.93\times10^{-7}$ |
| 4 | 15911116 | rs149191979 | G | A | 3.3% | 0.37 [0.21, 0.52] | $3.96\times10^{-6}$ | 0.21 [-0.10, 0.53] | 0.182 | 0.34 [0.20, 0.48] | $2.06\times10^{-6}$ |
| 5 | 8037516 | rs139744704 | T | C | 1.5% | -0.62 [-0.87, -0.36] | $2.36\times10^{-6}$ | 0.35 [-0.29, 0.99] | 0.282 | -0.48 [-0.72, -0.25] | $6.85\times10^{-5}$ |
| 5 | 38101390 | 5:38101390 | A | G | 1.5% | 0.60 [0.34, 0.87] | $6.10\times10^{-6}$ | -0.16 [-0.65, 0.33] | 0.522 | 0.43 [0.20, 0.67] | $2.52\times10^{-4}$ |
| 5 | 77235124 | rs78875199 | C | T | 1.8% | -0.60 [-0.83, -0.37] | $4.68\times10^{-7}$ | -0.11 [-0.50, 0.29] | 0.591 | -0.47 [-0.67, -0.27] | $3.99\times10^{-6}$ |
| 5 | 133553870 | rs76623936 | T | C | 1.7% | -0.43 [-0.62, -0.24] | $5.15\times10^{-6}$ | -0.05 [-0.50, 0.40] | 0.817 | -0.37 [-0.55, -0.20] | $1.90\times10^{-5}$ |

| | | | | | | | | | | | |
|---|---|---|---|---|---|---|---|---|---|---|---|
| 5 | 138949362 | rs261532 | G | T | 29.7% | 0.11 [0.06, 0.16] | $5.47×10^{-6}$ | -0.10 [-0.21, 0.02] | 0.094 | 0.08 [0.04, 0.12] | $3.91×10^{-4}$ |
| 8 | 104121210 | rs10111377 | A | G | 38.3% | 0.11 [0.06, 0.16] | $4.46×10^{-6}$ | 0.11 [0.00, 0.22] | 0.054 | 0.11 [0.07, 0.15] | $4.75×10^{-7}$ |
| 9 | 103502585 | rs10989235 | G | A | 8.4% | -0.23 [-0.33, -0.13] | $6.94×10^{-6}$ | -0.12 [-0.30, 0.07] | 0.224 | -0.20 [-0.29, -0.12] | $4.38×10^{-6}$ |
| 11 | 125945244 | rs142022403 | T | C | 2.2% | -0.50 [-0.71, -0.28] | $6.52×10^{-6}$ | 0.30 [-0.03, 0.64] | 0.079 | -0.26 [-0.45, -0.08] | 0.005 |
| 12 | 125685296 | rs35050183 | T | C | 39.5% | -0.12 [-0.18, -0.07] | $9.84×10^{-6}$ | -0.08 [-0.19, 0.03] | 0.151 | -0.11 [-0.16, -0.07] | $3.86×10^{-6}$ |
| 13 | 22321759 | rs76363177 | A | G | 1.7% | -0.52 [-0.75, -0.30] | $4.29×10^{-6}$ | -0.25 [-0.57, 0.08] | 0.144 | -0.44 [-0.62, -0.25] | $3.71×10^{-6}$ |
| 15 | 94002646 | rs149558104 | G | A | 1.7% | 0.57 [0.35, 0.80] | $8.55×10^{-7}$ | -0.34 [-0.78, 0.10] | 0.128 | 0.38 [0.18, 0.58] | $2.40×10^{-4}$ |
| 16 | 52507756 | rs141145965 | A | G | 1.8% | 0.48 [0.28, 0.69] | $5.02×10^{-6}$ | 0.31 [-0.21, 0.84] | 0.245 | 0.46 [0.27, 0.65] | $2.90×10^{-6}$ |
| 17 | 30589640 | rs147700400 | G | C | 1.5% | 0.81 [0.53, 1.09] | $1.18×10^{-8}$ | -0.22 [-0.55, 0.12] | 0.203 | 0.39 [0.17, 0.60] | $4.01×10^{-4}$ |
| 17 | 60499083 | rs144891349 | C | A | 5.6% | -0.28 [-0.41, -0.16] | $7.34×10^{-6}$ | 0.05 [-0.23, 0.34] | 0.712 | -0.23 [-0.34, -0.11] | $8.45×10^{-5}$ |
| 18 | 74794097 | rs2406710 | T | C | 97.3% | -0.39 [-0.56, -0.22] | $3.86×10^{-6}$ | -0.24 [-0.59, 0.11] | 0.184 | -0.36 [-0.51, -0.21] | $2.36×10^{-6}$ |
| 19 | 32840888 | rs74966282 | T | G | 1.6% | -0.50 [-0.72, -0.28] | $9.62×10^{-6}$ | 0.32 [-0.26, 0.91] | 0.283 | -0.40 [-0.61, -0.19] | $1.56×10^{-4}$ |
| 20 | 40894261 | rs6513775 | T | A | 45.0% | 0.13 [0.08, 0.18] | $1.83×10^{-6}$ | 0.09 [-0.02, 0.19] | 0.106 | 0.12 [0.07, 0.17] | $7.45×10^{-7}$ |
| 20 | 56516104 | rs6015146 | T | C | 97.8% | 0.48 [0.27, 0.69] | $8.53×10^{-6}$ | 0.00 [-0.39, 0.39] | 0.989 | 0.37 [0.19, 0.56] | $8.55×10^{-5}$ |
| 22 | 17479061 | rs147516555 | A | G | 2.1% | 0.50 [0.29, 0.70] | $1.89×10^{-6}$ | 0.17 [-0.28, 0.61] | 0.463 | 0.44 [0.25, 0.62] | $3.85×10^{-6}$ |
| 22 | 19833989 | rs1052988 | G | A | 23.8% | 0.14 [0.08, 0.20] | $6.01×10^{-6}$ | -0.06 [-0.18, 0.06] | 0.355 | 0.10 [0.05, 0.16] | $2.49×10^{-4}$ |

### Supplementary Table 2: Variants in credible set of signal on chromosome 1

Below is a table containing the seven variants on chromosome 1 in the credible set for the signal associated with FVC decline. The $r^2$ is the linkage disequilibrium of the variant with the sentinel variant rs115982800 (the variant with the strongest association with FVC decline). The effect estimates and p value are from the discovery GWAS. The "Posterior probability" column relates to the probability of the variant being causal as estimated by the Wakefield method. Annotation was taken from VEP[19]. Chr=Chromosome.

| Chr | Position | rsid | Effect allele | $r^2$ with sentinel variant | Posterior probability | Change in FVC (litres per year) [95% CI] | p | Annotation |
|---|---|---|---|---|---|---|---|---|
| 1 | 88741347 | rs115982800 | A | Sentinel | 25.7% | −0.14 [−0.19, −0.10] | $3.68\times10^{-10}$ | Intergenic |
| 1 | 88741980 | rs115590681 | T | 1.00 | 25.7% | −0.14 [−0.19, −0.10] | $3.72\times10^{-10}$ | Regulatory region (open chromatin) |
| 1 | 88743007 | rs116070595 | C | 1.00 | 25.7% | −0.14 [−0.19, −0.10] | $3.76\times10^{-10}$ | Intergenic |
| 1 | 88747734 | rs115194333 | T | 0.98 | 5.0% | −0.13 [−0.17, −0.09] | $3.48\times10^{-9}$ | Intergenic |
| 1 | 88756953 | rs76120044 | T | 0.97 | 5.0% | −0.13 [−0.17, −0.09] | $3.28\times10^{-9}$ | Intergenic |
| 1 | 88771866 | rs115304904 | C | 0.97 | 5.0% | −0.13 [−0.17, −0.09] | $3.14\times10^{-9}$ | Intergenic |
| 1 | 88759335 | rs143215745 | G | 0.97 | 3.6% | −0.13 [−0.17, −0.09] | $4.98\times10^{-9}$ | Intergenic |

**<u>Supplementary Table 3: Treatment interaction with rs115982800</u>**

Effect estimate from interaction term of *Treatment* × *SNP* × *Time*. I.e. a value below 0 would suggest that those with the rs115982800_A allele would have a worse treatment response (i.e. FVC would drop quicker) than those with the rs115982800_A allele not on the treatment.

| **Treatment** | **Effect estimate [95% CI] (Change in FVC ml/year)** | **p** |
|---|---|---|
| Nintedanib | 185 [−58, 428] | 0.136 |
| Pirfenidone | −108 [−303, 87] | 0.279 |
| Anti-microbial therapy | −56 [−246, 135] | 0.568 |

### Supplementary Table 4: Association of rs115982800 with other traits

[a] Effect sizes are with respect to the A allele of rs115982800 associated with declining FVC. [b] The general population lung function analysis was performed by regressing each SNP on extracted random time effects for each individual from a longitudinal model, meaning effect estimates were not available.

| Analysis | Sample size | Units | Effect estimate [a] [95% CI] | p |
|---|---|---|---|---|
| **i) Association with IPF traits** | | | | |
| Longitudinal FVC | 1,329 IPF cases | litres per year | −0.14 [−0.19, −0.10] | $3.68\times10^{-12}$ |
| Longitudinal DLco | 975 IPF cases | mmol/min/kPa/year | −0.09 [−0.20, 0.03] | 0.141 |
| IPF risk | 4,125 IPF cases vs 20,464 controls | Odds ratio | 0.91 [0.81, 1.03] | 0.142 |
| **ii) Association with lung function in general population** | | | | |
| Longitudinal $FEV_1$ | 32,013 general population (UK Biobank) | [b] | [b] | 0.690 |
| Longitudinal FVC | | [b] | [b] | 0.600 |
| Longitudinal $FEV_1$ | | [b] | [b] | 0.794 |
| **iii) Previously reported associations** | | | | |
| Dothiepin medication | 148 cases vs 361,141 controls (UK Biobank) | Odds ratio | 3.44 | $5.50\times10^{-6}$ |

**Supplementary Table 5: Gene prioritisation analyses using phenotype associations**

As part of the gene prioritisation analysis for the signal associated with FVC decline, we investigated whether nearby genes were associated with relevant phenotypes. Firstly, Mendelian diseases associated with rare genetic changes in nearby genes were investigated. Secondly, phenotypes associated with rare variants in these genes (either through single variant tests or gene-based tests) in the AZ PheWAS Portal. Finally, phenotypes recorded when the gene is knocked out in mouse models. Relevant phenotypes are shown in bold in red and potentially relevant phenotypes are shown in bold in orange. Genes not yet included in the International Mouse Phenotype Consortium are denoted with NA.

| | | | | Mendelian diseases | | AZ PheWAS Portal Associations | | Mouse knockout phenotypes |
|---|---|---|---|---|---|---|---|---|
| Chr | Start | End | Gene | Orphanet | OMIM | Single Variant | Gene | |
| 1 | 85715636 | 85725355 | C1orf52 | | | | | NA |
| 1 | 85731459 | 85742587 | *BCL10* | MALT lymphoma, Severe immunodeficiency, BENTA disease, Polyclonal B-cell lymphocytosis | Immunodeficiency, MALT Lymphoma, Follicula lymphoma, male germ cell tumor, mesothelioma, Sezary syndrome | | | NA |
| 1 | 85742040 | 85743771 | *LOC646626* | | | | | NA |
| 1 | 85784167 | 86044046 | *DDAH1* | | | Standing height | | erythrocyte cell number, grip strength |
| 1 | 86046443 | 86049648 | *CYR61* | Congenital heart defect (diagnostic test) | | Impedance of arm, leg and body | | NA |
| 1 | 86115105 | 86174116 | *ZNHIT6* | Autism | | | | enlarged lympth nodes, hemorrhage, abnormal lymph node morphology, preweaning lethality, prepulse inhibition |
| 1 | 86194915 | 86622121 | *COL24A1* | | | 6mm strong/weak merdiain (eye) | | body length, grip strength |
| 1 | 86812506 | 86862025 | *ODF2L* | Intellectual disability | | Basal metabolic rate, leg fat free mass, leg predicted mass, standing height, trunk fat free mass, trunk predicted mass, whole body fat free mass, whole body water mass | | small kidney, blind uterus, abnormal kidney morphology |
| 1 | 86823314 | 86823370 | *MIR7856* | | | | | NA |
| 1 | 86889768 | 86922240 | *CLCA2* | | | | | NA |
| 1 | 86934525 | 86965974 | *CLCA1* | | | | | hydrometra, abnormal vibrissa morphology, small superior vagus ganglion |
| 1 | 87012758 | 87046432 | *CLCA4* | Hepatic diseases | | | | NA |

| | | | | | | | | |
|---|---|---|---|---|---|---|---|---|
| 1 | 87099958 | 87121059 | *CLCA3P* | | | | | NA |
| 1 | 87170252 | 87213867 | *SH3GLB1* | | | | | NA |
| 1 | 87328127 | 87380107 | *SEP15* | | | | | NA |
| 1 | 87380334 | 87575681 | *HS2ST1* | | Neurofacioskeletal syndrome with or without renal agenesis | | | NA |
| 1 | 87595447 | 87634886 | *LINC01140* | | | | | NA |
| 1 | 87678351 | 87717014 | *LOC101927844* | | | | | NA |
| 1 | 87794150 | 87814607 | *LMO4* | | | | **FVC**, **$FEV_1$** | NA |
| 1 | 87819209 | 87837338 | *LOC100505768* | | | | | NA |
| 1 | 89003195 | 89150887 | *LOC101927891* | | | | | NA |
| 1 | 89149921 | 89301938 | *PKN2* | **Autoinflammatory diseases (diagnostic panel - candidate)** | | corneal hysteresis, corneal resistance factor, Gamma glutamyltransferase, Aspartate aminotransferase, urea, sitting height, standing height, **% predicted $FEV_1$** | | decreased exploration, startle reflex, circulating bilirubin, circulating transferrin, preweaning lethality, abnormal retina morphology, circulating cholesterol, abnormal retina vasculature morphology, total circulating protein, circulating HDL |
| 1 | 89318320 | 89357301 | *GTF2B* | | | | | embryonic lethality, preweaning lethality |
| 1 | 89401455 | 89458643 | *CCBL2* | | | Gamma glutamyltransferase, standing height, corneal resistance factor, Intra-ocular pressure\| Goldmann-correlated (left), urea | | NA |
| 1 | 89445138 | 89458643 | *RBMXL1* | | | | | NA |
| 1 | 89472359 | 89488549 | *GBP3* | | | Inhalation anaesthetic using muscle relaxant | | |
| 1 | 89517986 | 89531043 | *GBP1* | | | | | NA |
| 1 | 89571815 | 89591842 | *GBP2* | | | | | prolonged RR interval, fasting glucose level, increased heart rate variability |
| 1 | 89597433 | 89641723 | *GBP7* | | | | | |
| 1 | 89646830 | 89664633 | *GBP4* | | | | | |
| 1 | 89724633 | 89738544 | *GBP5* | **Autoinflammatory diseases (diagnostic panel - candidate)** | | | | increased creatinine level |
| 1 | 89754940 | 89756045 | *LOC729930* | | | | | NA |
| 1 | 89829435 | 89853719 | *GBP6* | | | Inhalation anaesthetic using muscle relaxant | | NA |

| | | | | | | | | |
|---|---|---|---|---|---|---|---|---|
| 1 | 89873237 | 89890493 | *GBP1P1* | | | | | NA |
| 1 | 89990396 | 90063420 | *LRRC8B* | | | | | NA |
| 1 | 90090407 | 90098453 | *FLJ27354* | | | | | NA |
| 1 | 90098643 | 90185094 | *LRRC8C* | | | | | abnormal eye morphology, small testis, abnormal spleen morphology, abnormal testis morphology |
| 1 | 90286572 | 90401989 | *LRRC8D* | | | | | increased Alkaline phosphatase |
| 1 | 90458823 | 90460525 | *GEMIN8P4* | | | | | NA |
| 1 | 90460677 | 90494094 | *ZNF326* | | | | | NA |
| 1 | 91177578 | 91182794 | *BARHL2* | | | | | NA |
| 1 | 91380856 | 91487812 | *ZNF644* | | Myopia 21 | | | abnormal uterus morphology, increased circulating potassium, abnormal skin morphology, increased blood urea nitrogen, decreased body weight, abnormal testis morphology, decreased mean corpuscular volume, increased circulating HDL, decreased mean corpuscular heamoglobin, enlarged uterus, increased circulating alkaline phosphatse, decreased circulating chloride |
| 1 | 91726322 | 91870426 | *HFM1* | Ovarian failure/insufficiency, Sex development disorders, Female infertility | Premature ovarian failure | **IGF-1** | | NA |

**Supplementary Table 6: Association with FVC and DLco decline for variants previously reported as associated with IPF risk**

Below is a table showing the association between the 19 IPF risk variants reported in Allen et al (2021)[20] and FVC and DLco decline. The effect allele is the allele associated with increased risk of IPF. Chr=Chromosome. EAF=Effect allele frequency. OR=Odds ratio. CI=Confidence interval.

| Chr | Position | rsid | Locus | Ref allele | Effect allele | EAF | Risk | | FVC | | DLco | |
|---|---|---|---|---|---|---|---|---|---|---|---|---|
| | | | | | | | **OR [95% CI]** | **p** | **β [95% CI]** | **p** | **β [95% CI]** | **p** |
| 3 | 44903434 | rs2292181 | *KIF15* | G | C | 5.2% | 1.52 [1.36, 1.70] | $3.95\times10^{-13}$ | 0.00 [−0.04, 0.04] | 0.935 | −0.09 [−0.19, 0.01] | 0.070 |
| 3 | 169486271 | rs9860874 | *TERC* | C | A | 27.6% | 1.29 [1.22, 1.37] | $6.49\times10^{-18}$ | 0.01 [−0.01, 0.03] | 0.411 | −0.02 [−0.07, 0.04] | 0.514 |
| 4 | 89837808 | rs2609259 | *FAM13A* | C | A | 22.4% | 1.30 [1.22, 1.39] | $6.47\times10^{-17}$ | 0.01 [−0.01, 0.04] | 0.351 | 0.03 [−0.02, 0.09] | 0.257 |
| 5 | 1282414 | rs7725218 | *TERT* | A | G | 67.1% | 1.41 [1.33, 1.50] | $4.90\times10^{-32}$ | −0.02 [−0.04, 0.01] | 0.191 | −0.04 [−0.10, 0.02] | 0.194 |
| 6 | 7563232 | rs2076295 | *DSP* | T | G | 46.7% | 1.49 [1.41, 1.57] | $1.50\times10^{-48}$ | −0.01 [−0.03, 0.01] | 0.507 | 0.03 [−0.03, 0.08] | 0.363 |
| 7 | 1868761 | rs12537430 | *MAD1L1* | A | G | 62.5% | 1.28 [1.21, 1.35] | $4.20\times10^{-18}$ | −0.01 [−0.04, 0.02] | 0.470 | 0.00 [−0.06, 0.06] | 0.961 |
| 7 | 99630342 | rs2897075 | *ZKSCAN1* | C | T | 38.2% | 1.30 [1.23, 1.37] | $1.77\times10^{-21}$ | 0.00 [−0.02, 0.03] | 0.639 | 0.05 [−0.01, 0.10] | 0.096 |
| 8 | 120940206 | rs10808505 | *DEPTOR* | G | T | 57.3% | 1.20 [1.13, 1.26] | $6.03\times10^{-11}$ | 0.00 [−0.03, 0.02] | 0.717 | 0.00 [−0.06, 0.05] | 0.967 |
| 10 | 111229861 | rs79684490 | 10q25.1 | G | A | 4.6% | 1.40 [1.24, 1.57] | $3.52\times10^{-8}$ | 0.03 [−0.02, 0.07] | 0.262 | 0.01 [−0.10, 0.13] | 0.837 |
| 11 | 1241221 | rs35705950 | *MUC5B* | G | T | 14.5% | 5.06 [4.69, 5.47] | $9.09\times10^{-418}$ | 0.02 [−0.01, 0.04] | 0.251 | −0.01 [−0.07, 0.05] | 0.739 |
| 13 | 113534984 | rs9577395 | *ATP11A* | G | C | 79.1% | 1.29 [1.21, 1.38] | $4.78\times10^{-14}$ | −0.01 [−0.04, 0.02] | 0.479 | 0.01 [−0.06, 0.08] | 0.700 |
| 15 | 40716253 | rs2304645 | *IVD* | G | C | 52.6% | 1.28 [1.21, 1.35] | $8.66\times10^{-20}$ | −0.01 [−0.04, 0.01] | 0.190 | −0.03 [−0.07, 0.02] | 0.329 |
| 15 | 40931708 | rs12912339 | *KNL1* | G | A | 15.9% | 1.30 [1.21, 1.39] | $7.41\times10^{-13}$ | 0.01 [−0.01, 0.04] | 0.301 | −0.06 [−0.13, 0.00] | 0.067 |
| 15 | 86287910 | rs11073517 | *AKAP13* | C | T | 32.7% | 1.19 [1.13, 1.26] | $1.36\times10^{-9}$ | 0.02 [0.00, 0.05] | 0.037 | 0.00 [−0.05, 0.06] | 0.939 |
| 16 | 162240 | rs74614704 | *NPRL3* | G | A | 5.6% | 1.49 [1.33, 1.67] | $2.57\times10^{-12}$ | −0.01 [−0.05, 0.03] | 0.546 | −0.06 [−0.16, 0.04] | 0.231 |
| 17 | 44214888 | rs2077551 | 17q21.31 | C | T | 80.7% | 1.42 [1.32, 1.53] | $1.92\times10^{-20}$ | - | - | - | - |
| 19 | 4717672 | rs12610495 | *DPP9* | A | G | 30.6% | 1.28 [1.21, 1.36] | $2.58\times10^{-16}$ | 0.01 [−0.01, 0.03] | 0.348 | 0.04 [−0.02, 0.09] | 0.158 |
| 20 | 62284170 | rs112087793 | *STMN3* | T | C | 91.5% | 1.34 [1.21, 1.48] | $1.09\times10^{-8}$ | 0.02 [−0.03, 0.06] | 0.478 | −0.01 [−0.12, 0.10] | 0.796 |
| 20 | 62324391 | rs41308092 | *RTEL1* | G | A | 2.1% | 1.75 [1.45, 2.10] | $3.13\times10^{-9}$ | 0.00 [−0.07, 0.07] | 0.910 | 0.02 [−0.15, 0.18] | 0.826 |

**Supplementary Table 7: Genes showing suggestive association ($p<10^{-5}$) with FVC or DLco decline as part of gene-based test**

Gene-based tests were performed using Vegas2 to combine the effect of multiple variants. Chr=Chromosome. i) Genes with a suggestive significant association with FVC decline. No variants reached a Bonferroni corrected threshold of $2.28\times10^{-6}$ was used for significance (based on 21,910 genes included in the FVC analysis). ii) Genes with a suggestive significant association with DLco decline. No variants reached a Bonferroni corrected threshold of $2.28\times10^{-6}$ was used for significance (based on 21,911 genes included in the DLco analysis).

**i) FVC**

| Gene | Chr | Position | Number of SNPs | p |
|---|---|---|---|---|
| *TMEM105* | 17 | 79285071-79304474 | 68 | $2.99\times10^{-6}$ |

**ii) DLco**

| Gene | Chr | Position | Number of SNPs | p |
|---|---|---|---|---|
| *BDH2* | 4 | 103998781-104021024 | 49 | $9.99\times10^{-6}$ |

**Supplementary Table 8: Genes in the pathway "GO:0051302_regulation_of_cell_division" showing marginal association with DLco decline**

| Genes is GO:0051302_regulation_of_cell_division | Empirical p value for enrichment of FVC decline | Empirical p value for enrichment of DLco decline |
|---|---|---|
| *CCNL2, KLHL21, MAD2L2, TAS1R2, EIF4G3, WNT4, CEP85, GPR3, RCC1, YBX1, CDC20, PLK3, STIL, SH3GLB1, PKN2, CDC14A, PSRC1, PROK1, TXNIP, HORMAD1, TPR, ASPM, KIF14, NEK2, PROX1, CENPF, TGFB2, CCSAP, MAP10, ZBTB18, ADAM17, GEN1, ATRAID, SPDYA, BIRC6, PRKCE, MSH2, GKN1, TGFA, RNF103-CHMP3, BUB1, ANAPC1, IL1B, CCNT2, PRPF40A, PKP4, CD28, OBSL1, CUL3, HTR2B, DAZL, UBE2E1, DYNC1LI1, PDCD6IP, NME6, RHOA, WNT5A, CHMP2B, DRD3, PIK3R4, CCNL1, ECT2, TP63, FGFR3, MSX1, ANAPC4, PTTG2, BTC, FGF5, PKD2, CENPE, EGF, MAD2L1, FGF2, INTU, ANAPC10, SFRP2, MAP9, PDGFC, VEGFC, PRDM9, RXFP3, RAD1, FGF10, CCNB1, THBS4, CCNH, APC, UBE2B, NEUROG1, CDC25C, FGF1, PDGFRB, PTTG1, DUSP1, MSX2, GNB2L1, EDN1, TNF, PIM1, CCND3, VEGFA, KHDC3L, PHIP, TTK, CCNC, FBXO5, RPS6KA2, DLL1, PDGFA, MAD1L1, MACC1, HOXA13, ANLN, CDK13, CAMK2B, EGFR, CAV2, PTN, WEE2, SHH, TNKS, PIWIL2, NKX3-1, SOX17, MOS, CSPP1, TERF1, CHMP4C, FBXO43, MTBP, NSMCE2, MYC, ZNF16, DMRT1, CHMP5, SMC5, PTCH1, CDC26, DAB2IP, NEK6, SDCCAG3, ANAPC2, SVIL, CCNY, UBE2D1, STOX1, ANAPC16, C10orf99, PTEN, KIF20B, KIF11, FGF8, FGFR2, BUB3, INS, E2F8, NELL1, PAX6, CAT, MDK, VEGFB, CDCA5, LRP5, FGF3, ANAPC15, UVRAG, PDGFD, ATM, ZW10, CXCR5, CHEK1, FGF6, PHB2, TAS2R13, CDKN1B, PDE3A, CCNT1, RACGAP1, ESPL1, E2F7, USP44, IGF1, ANAPC7, CIT, ANAPC5, KNTC1, FGF9, BRCA2, RGCC, RB1, BORA, PCID2, CDC16, PRMT5, BMP4, DLGAP5, HSPA2, PGF, TGFB3, CALM1, CCNK, XRCC3, AKT1, BUB1B, ZFYVE19, NUSAP1, FGF7, KIF23, BBS4, BLM, IGF1R, SSTR5, PKD1, PKMYT1, CCP110, PLK1, LCMT1, SH2B1, CSNK2A2, VPS4A, PRDM7, OR1A2, AURKB, CENPV, FLCN, TOM1L2, GIT1, ZNF207, CDC6, BECN1, KIF18B, CDC27, PPP1R9B, TOM1L1, TEX14, PRKAR1A, BIRC5, ANAPC11, CHMP1B, PSMG2, PIK3C3, VPS4B, DAPK3, INSR, PIN1, CALR, GIPC1, SIRT2, TGFB1, CALM3, AURKC, CHMP2A, CSNK2A1, CDC25B, PLCB1, KIF3B, CHMP4B, SRC, L3MBTL1, UBE2C, NCOA3, AURKA, BMP7, EDN3, RANBP1, KLHL22, SUSD2, OSM, PDXP, PDGFB* | 0.219 | $8.80\times10^{-5}$ |

### Supplementary Figures

### Supplementary Figure 1: Quality control flowchart

#### i) Quality control for discovery genome-wide analyses

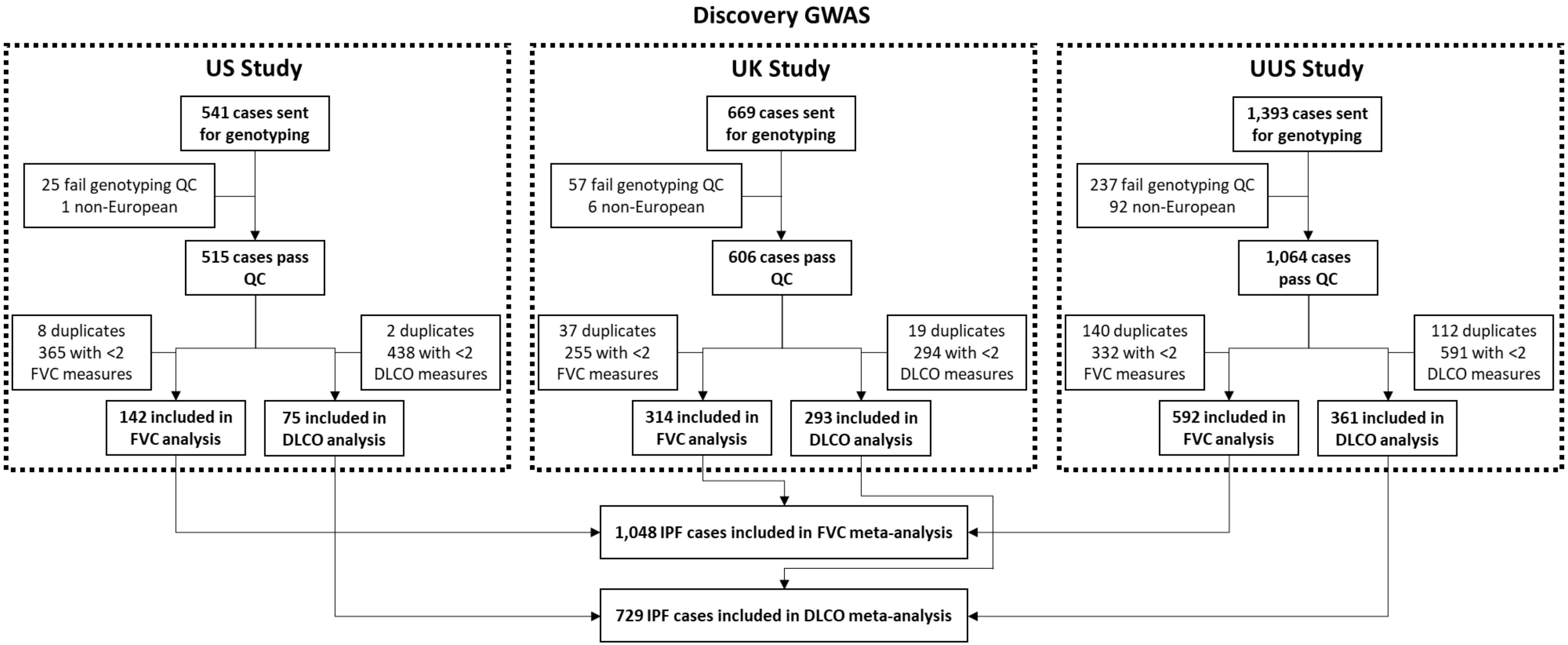

**ii) Quality control for follow-up analysis**

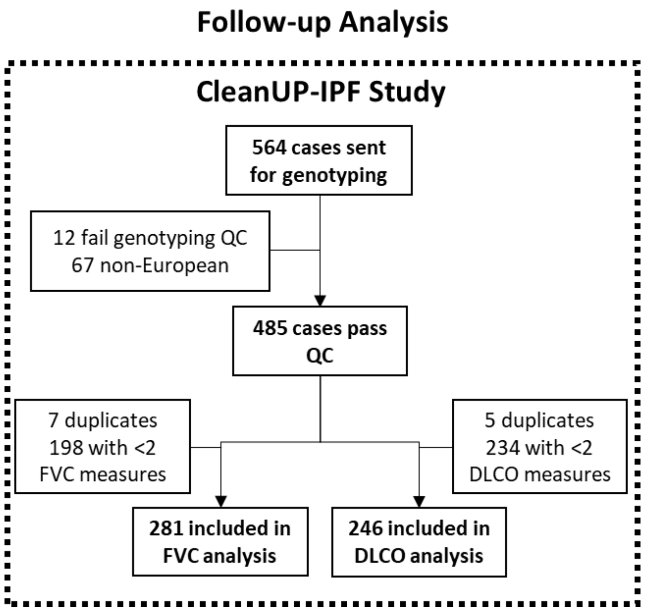

**<u>Supplementary Figure 2: Plots of longitudinal FVC and DLco over time</u>**

Each line shows an individual's change in FVC or DLco over time. Individuals are coloured by which study centre they were recruited.

i) **Longitudinal FVC and DLco for individuals from the US Study**

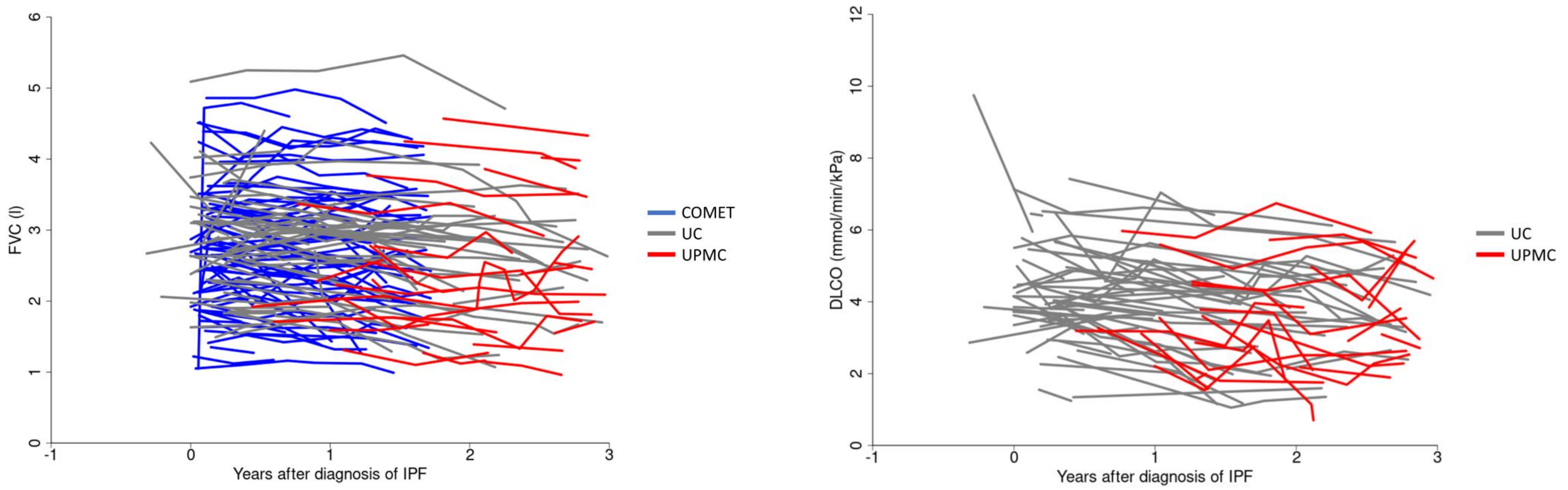

**ii) Longitudinal FVC and DLco for individuals from the UK Study**

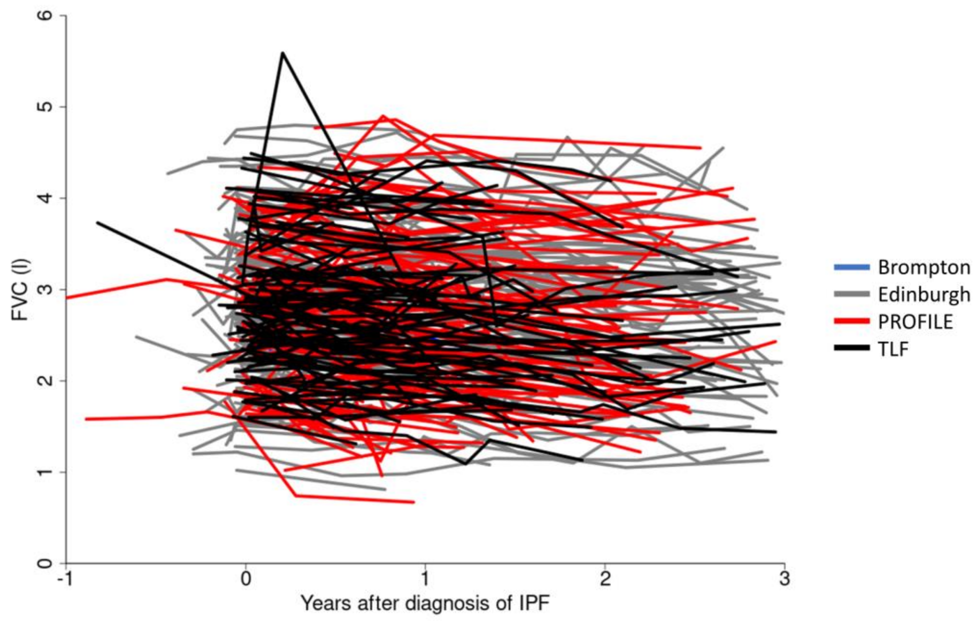

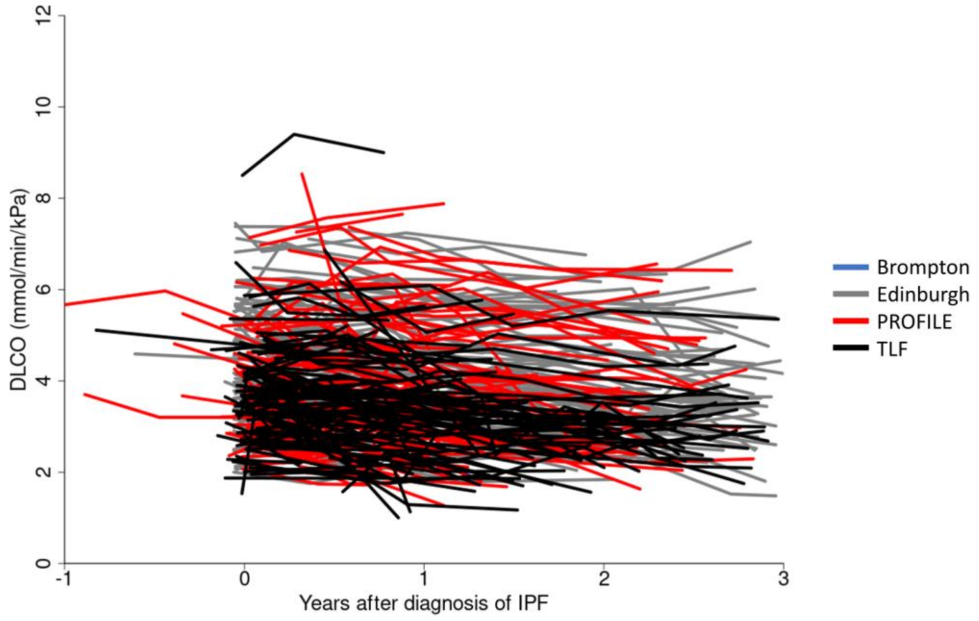

**iii) Longitudinal FVC and DLco for individuals from the UUS Study**

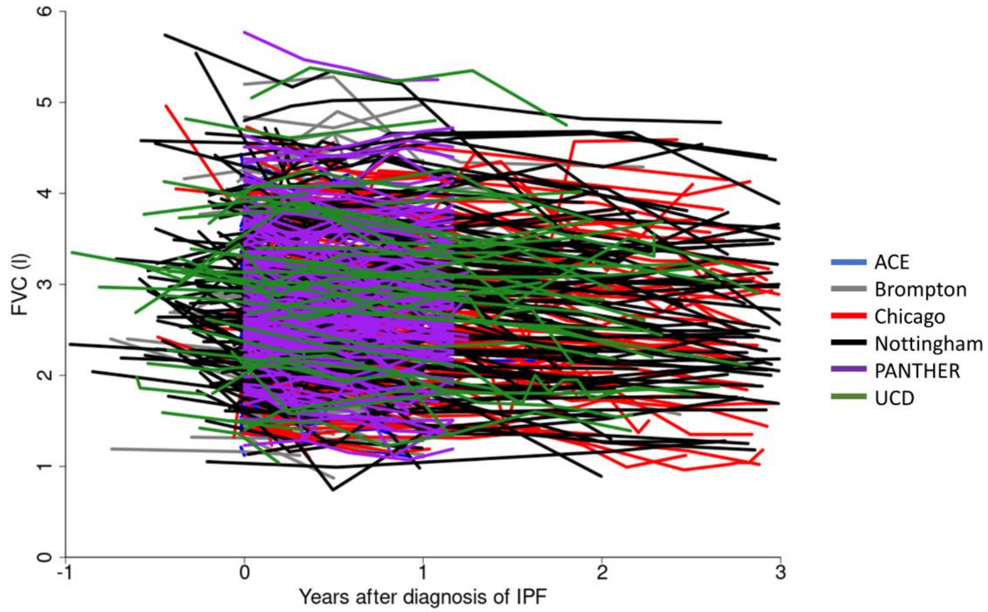

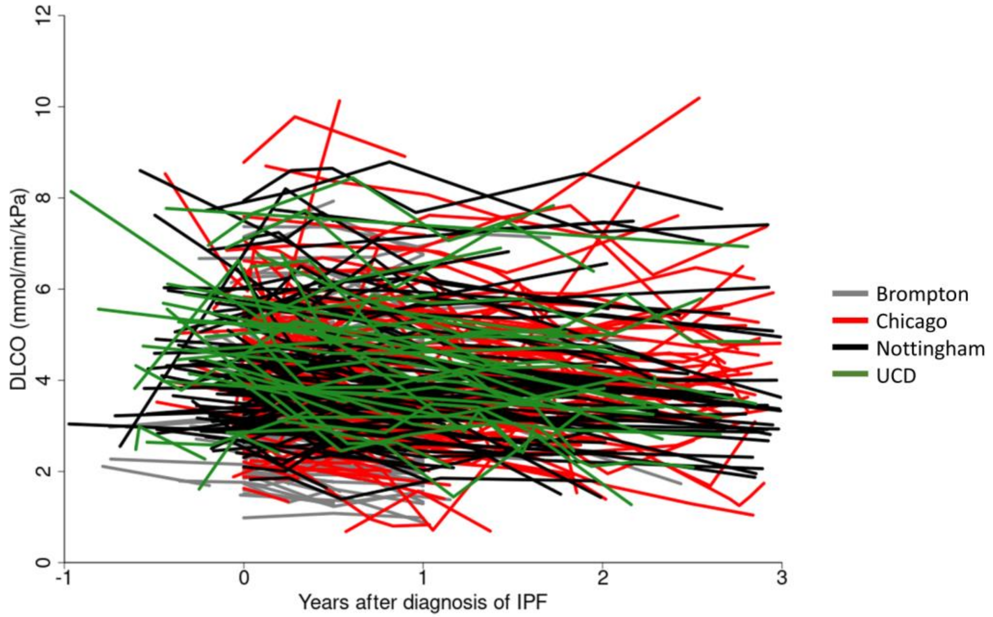

**iv) Longitudinal FVC and DLco for individuals from the CleanUP-IPF study**

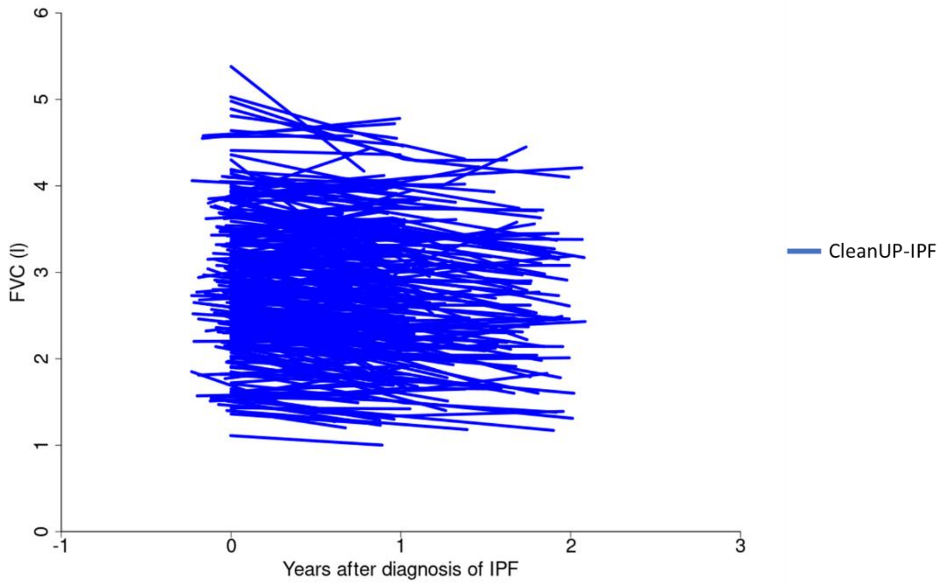

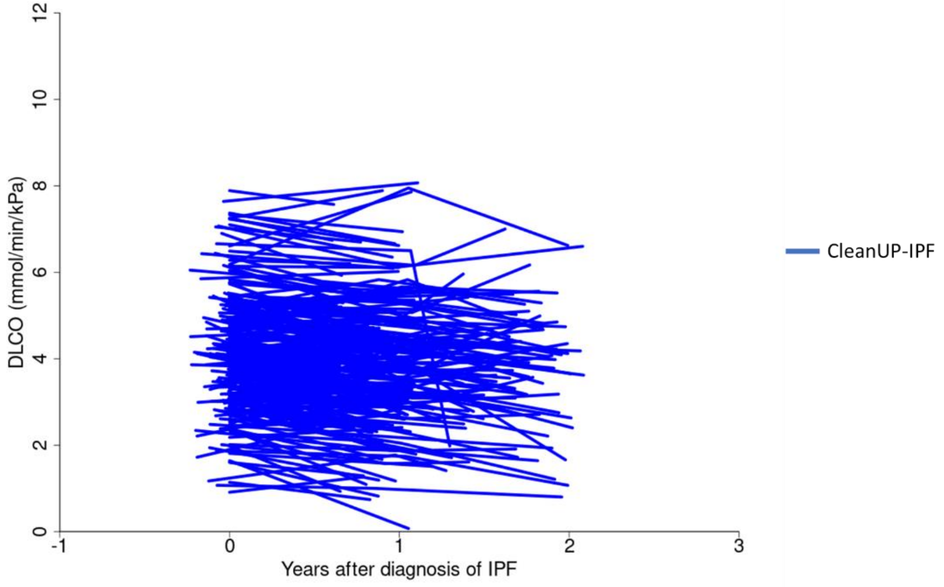

**Supplementary Figure 3: Histograms of the number of measures for FVC and DLco for each individual by study centre**

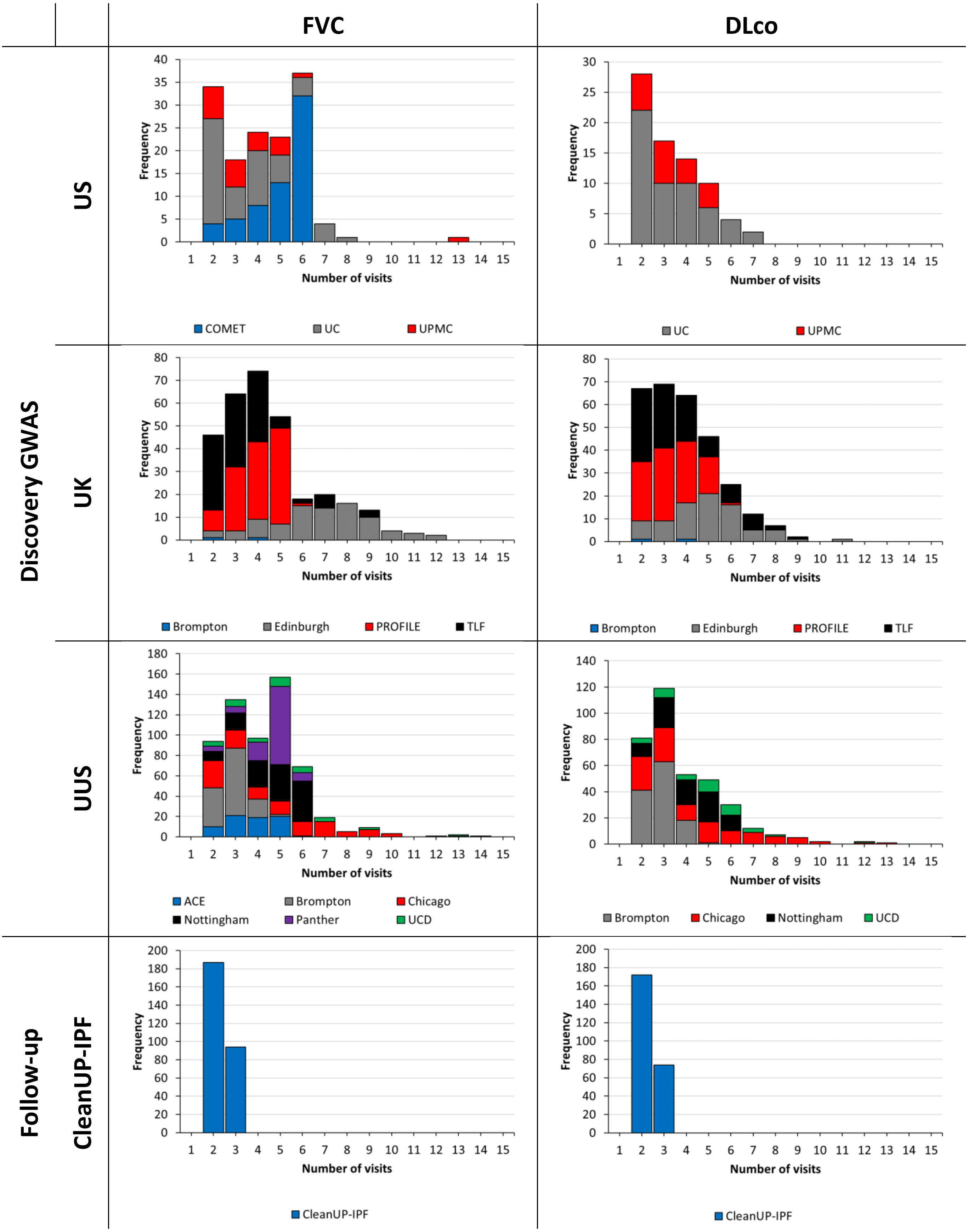

**Supplementary Figure 4: QQ plots for genome-wide longitudinal FVC and DLco analyses**

The QQ plot shows the expected distribution of –log(p values) against the observed distribution of –log(p values). The red lines show where these are equal.

**i) FVC**

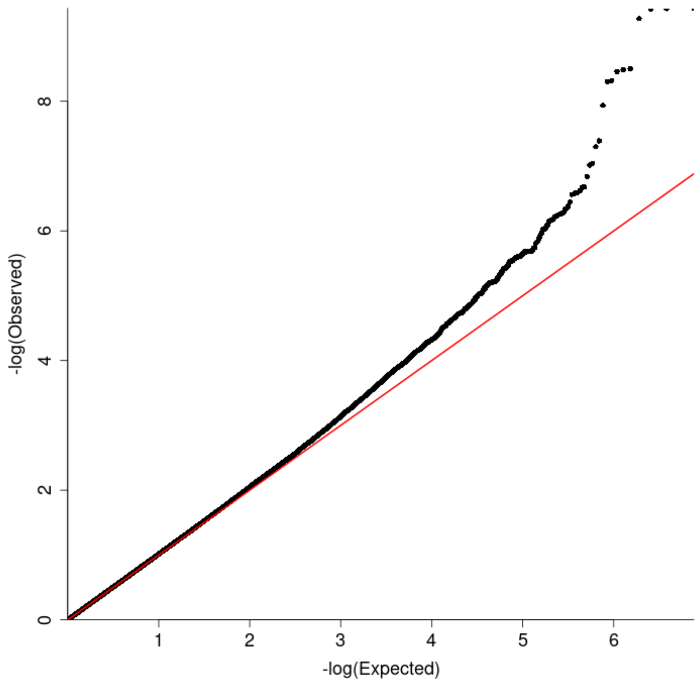

**ii) DLco**

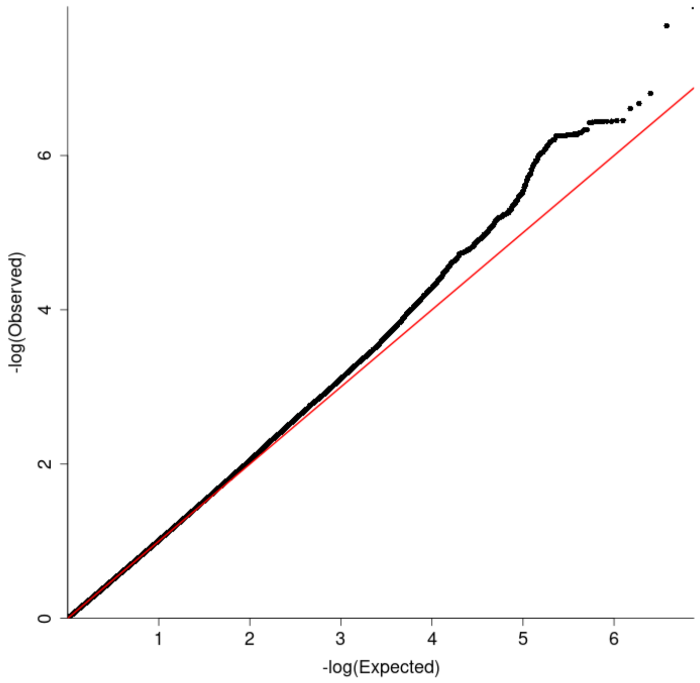

### Supplementary Figure 5: Comparison of FVC and DLco analysis results

#### i) Comparison of $-\log_{10}$(p values) from FVC and DLco analyses

Each point represents a genetic variant with its –log(p value) from the FVC meta-analysis on the x axis and the –log(p value) from the DLco meta-analysis on the y axis. The red lines show the genome-wide significance threshold of $p=5\times10^{-8}$.

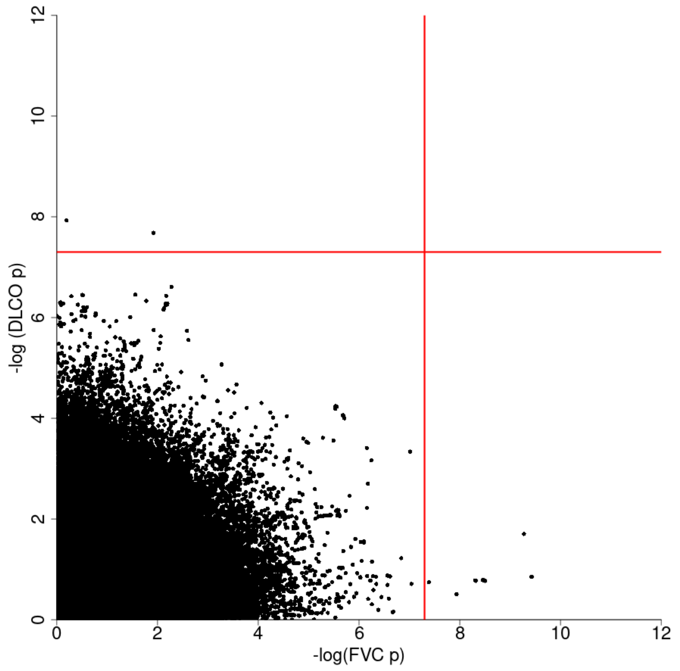

#### ii) Comparison of effect sizes from FVC and DLco analyses

Each point represents a genetic variant with the β from the FVC meta-analysis on the x axis and the β from the DLco meta-analysis on the y axis.

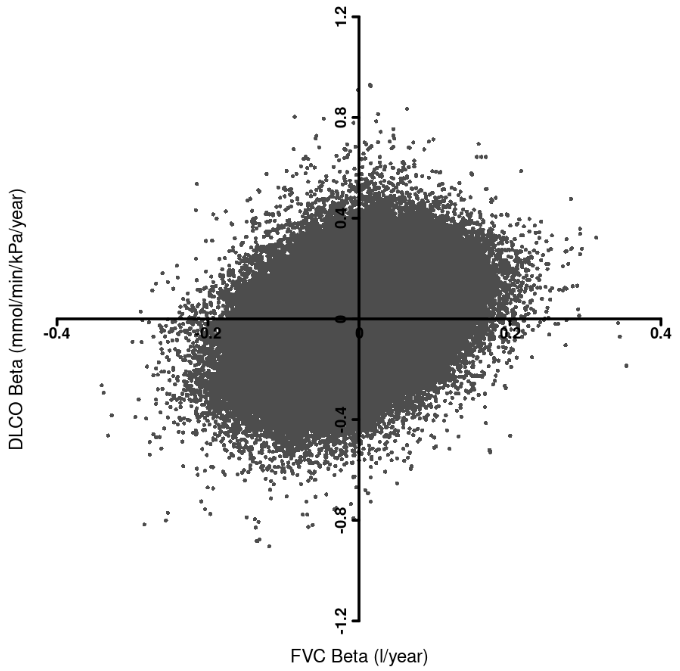

**Supplementary Figure 6: Longitudinal FVC coloured by rs115982800 genotype**

Each line shows an individual's FVC trajectory over time. Individuals with two copies of the reference allele G are shown in black, individuals with a GA genotype are shown in red and individuals with two copies of the risk allele (i.e. an AA genotype) are shown in blue.

**UK**

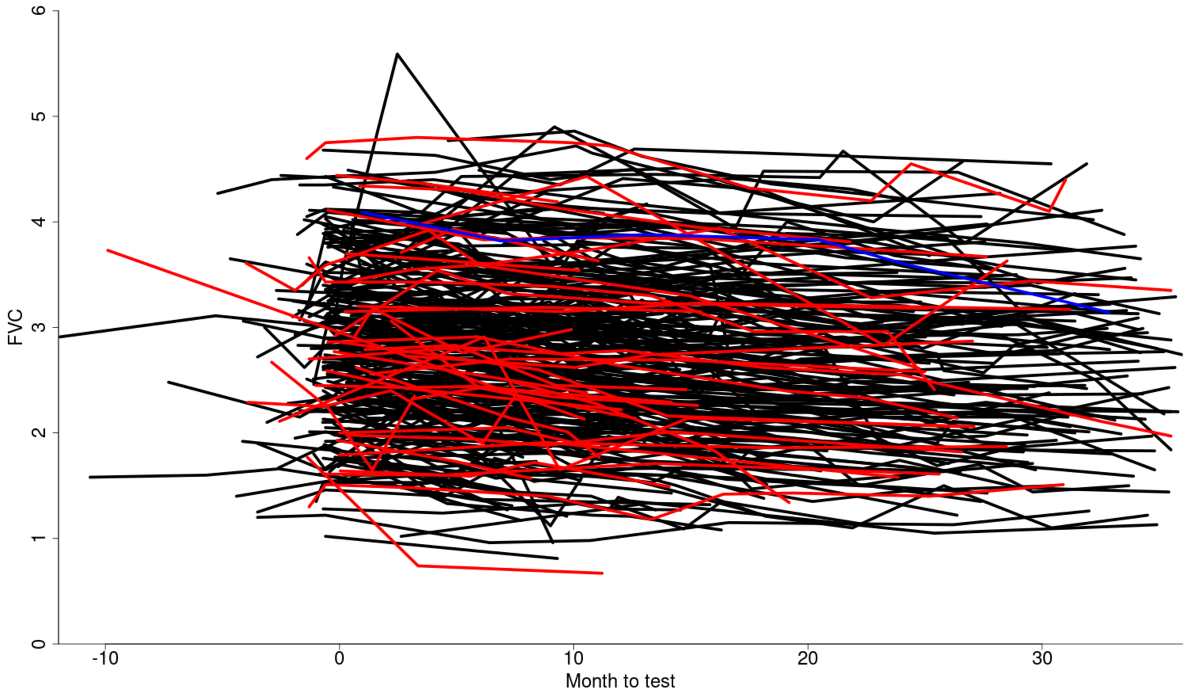

**UUS**

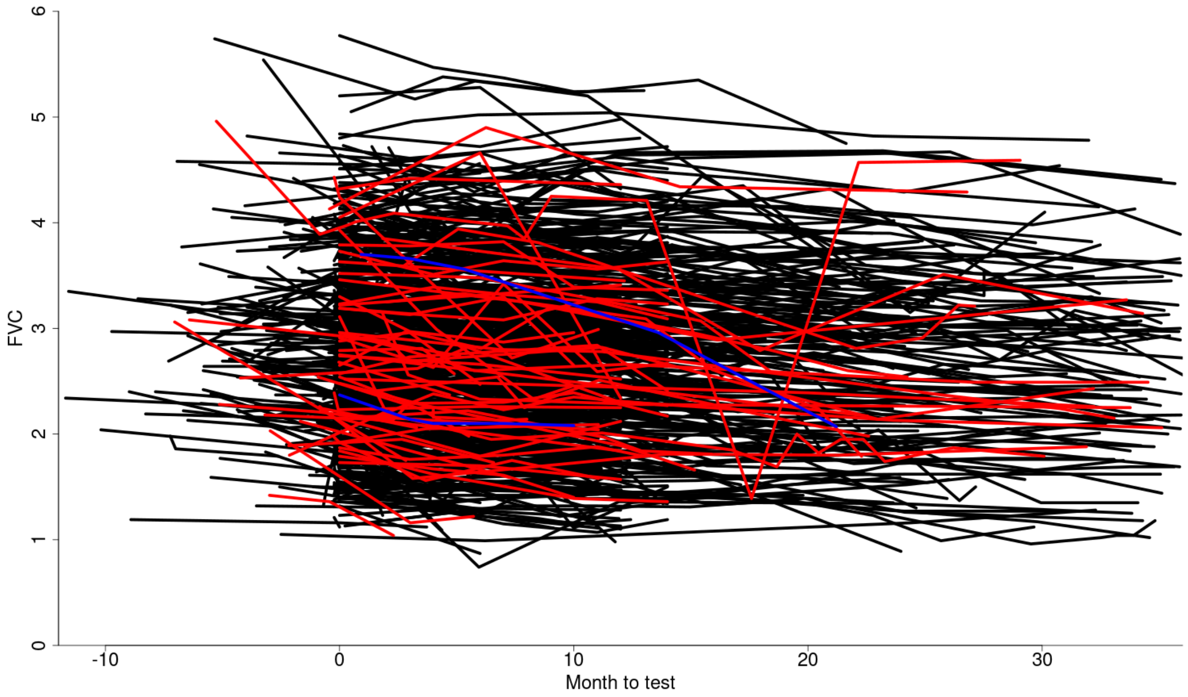

**US**

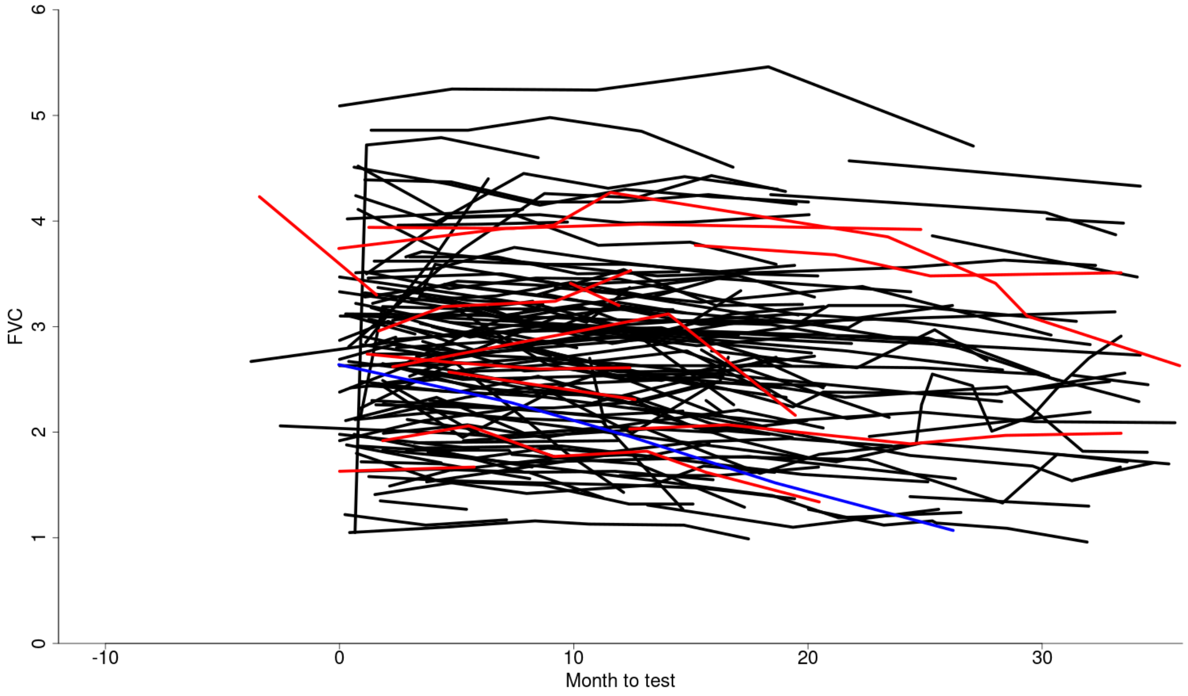

**CleanUP-IPF**

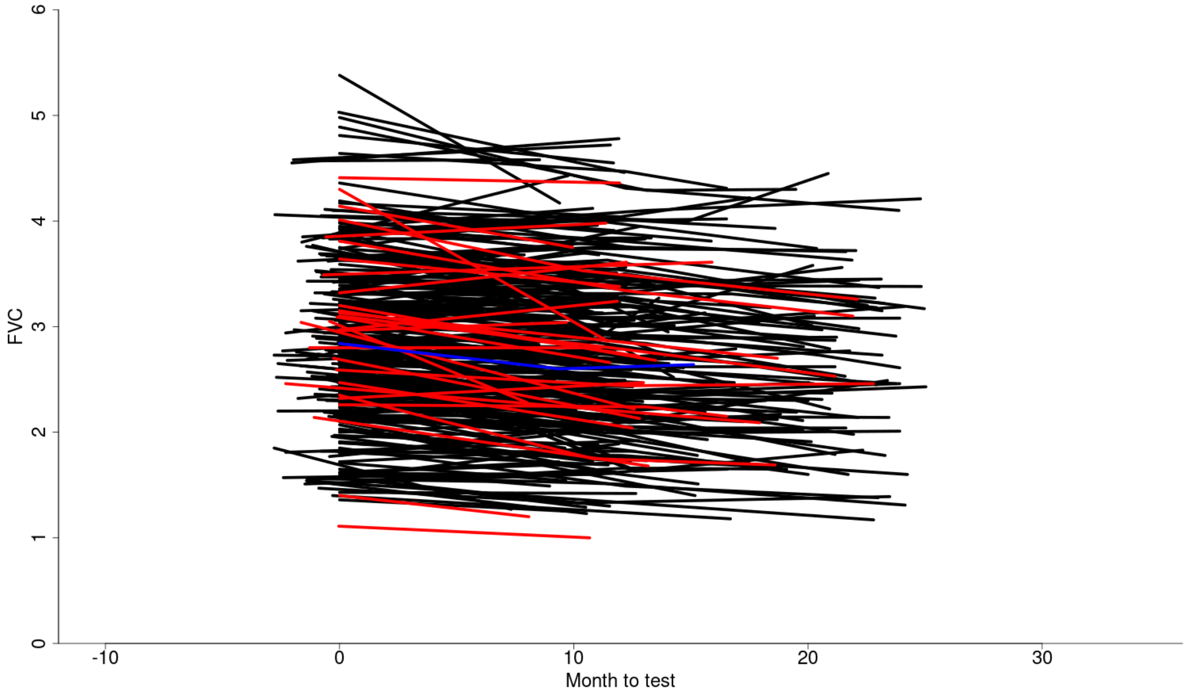

### Supplementary Figure 7: Forest plots for sensitivity analyses of rs115982800

Below are forest plots from the sensitivity analyses for rs115982800. The square in the middle gives the point estimate from each study and the lines show the 95% confidence intervals. The size of the box is relative to the weight given to the study in the meta-analysis (which is based on the inverse of the standard error).

#### i) Longitudinal models

Points to the left of the horizontal line suggest the variant is associated with decreasing FVC. The original analysis for rs115982800 is shown in black for each study. The sensitivity analysis restricting data to within 1 year of diagnosis is shown in blue. These give similar results to the original analysis for the UK and UUS studies, however the confidence intervals are wider due to the reduced power from reducing the number of measurements included in the analysis. The mixed linear models did not converge for the US or CleanUP-IPF studies due to there not being enough measurements, and therefore there are no lines for these studies. The red lines show the results from the longitudinal analysis when included as part of a joint model. For the UK, UUS and US study the variant becomes non-significant when incorporating drop-out due to death in the model. There was very little drop-out in the CleanUP-IPF study and therefore the results in the joint model are similar to that from the original genome-wide linear mixed model.

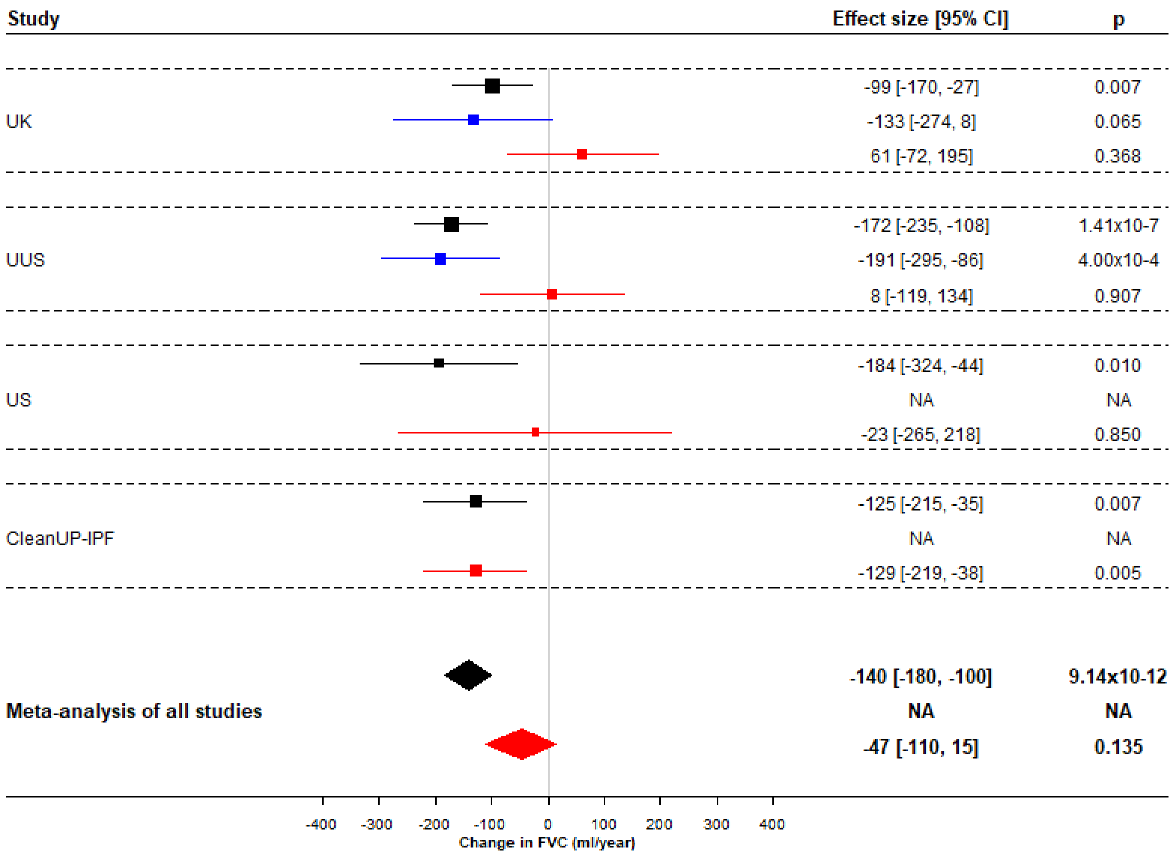

#### ii) Clinical model

Results when defining individuals as progressive if they had a decrease of FVC ≥ 10% in the first year or died. Results are given as odds ratios and plotted on a log scale. Points to the right of the line suggest the variant is associated with increased odds of being "progressive".

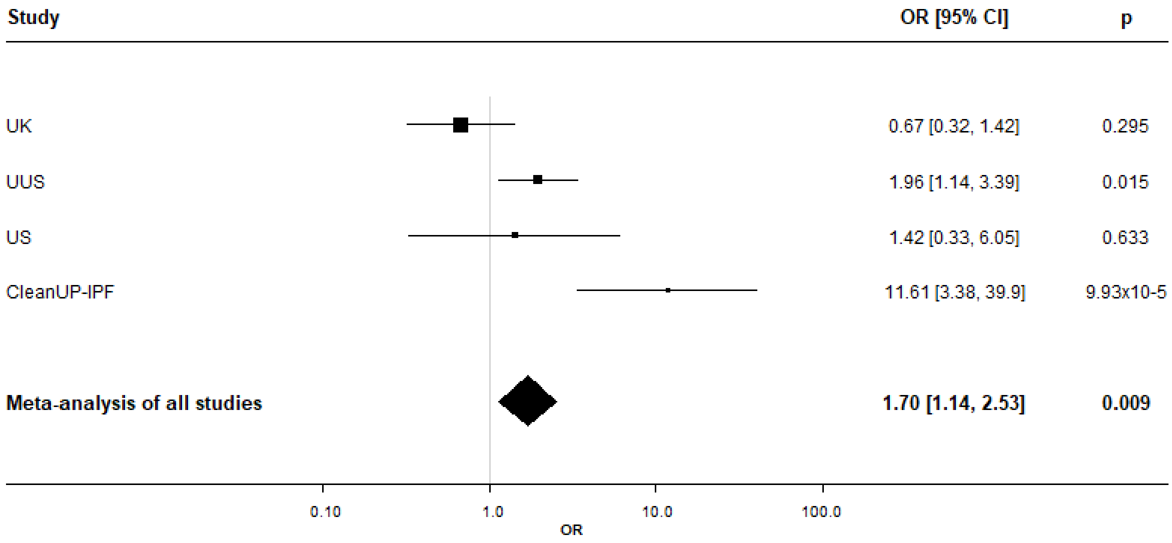

#### iii) Non-linear model

Results when including random effects of quadratic time and a quadratic time-by-SNP interaction term. The forest plot on the left shows the interaction between rs115982800 and linear time and the plot on the right shows the interaction between rs115982800 and quadratic time. These estimates come from the same model. The model did not converge when using the CleanUP-IPF study. Note: The quadratic effect size estimates are much smaller and are plotted on a different scale to the linear interaction terms.

**Linear time interaction term**

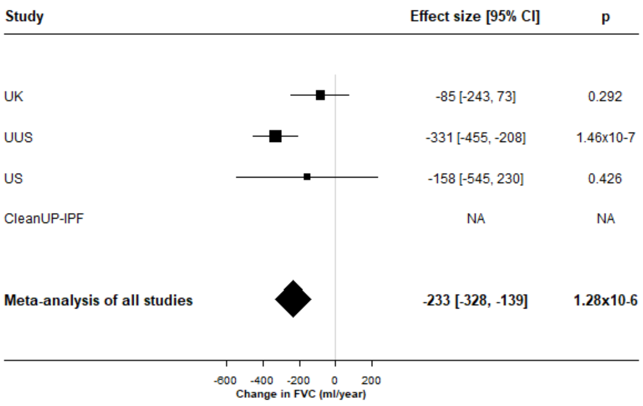

**Quadratic time interaction term**

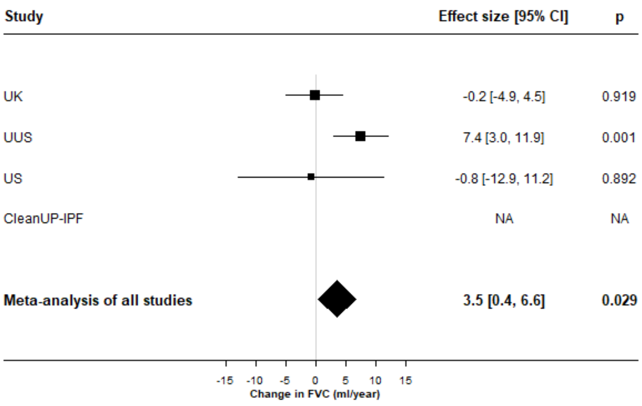

#### iv) Baseline lung function

Results when testing whether the variant rs115982800 was associated with the first measure of FVC (shown in black) or DLco (shown in blue). Points to the right of the vertical line suggest the variant would be associated with higher values of FVC or DLco at first visit.

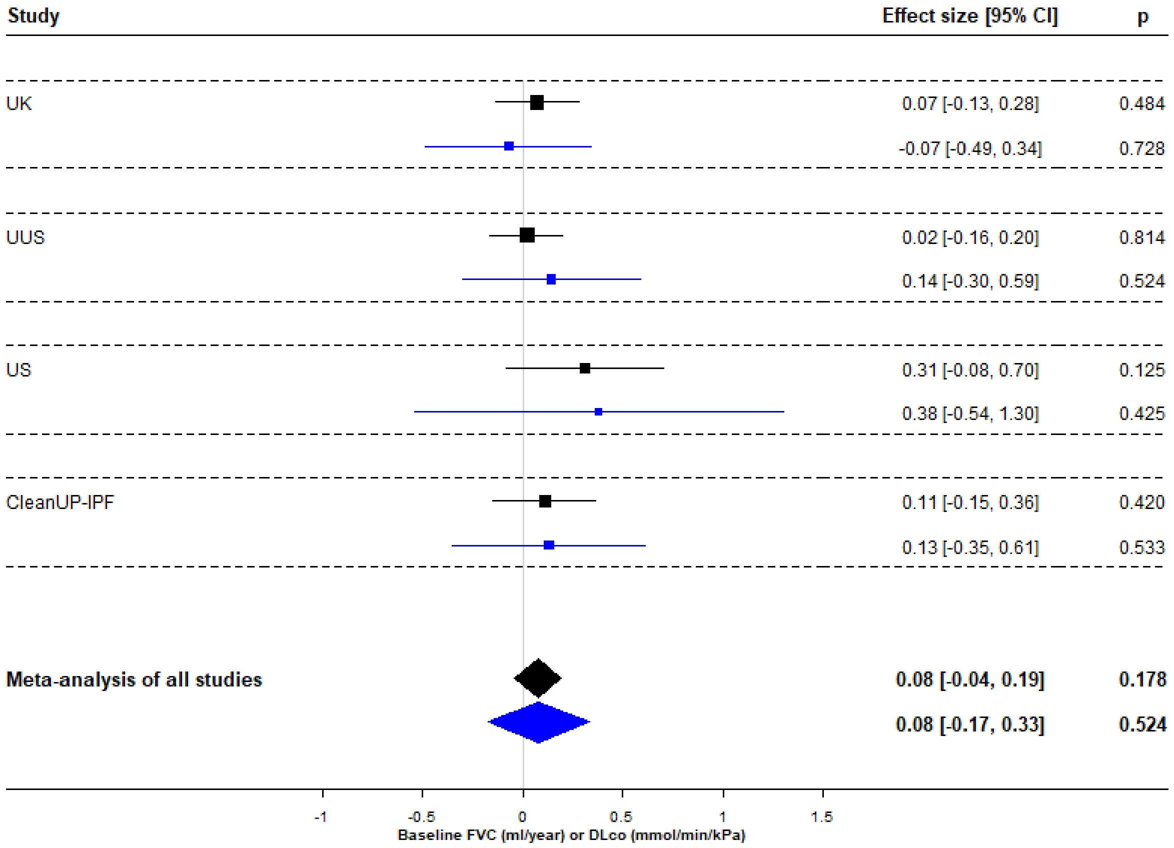

#### v) Age at baseline

Results when testing whether the variant rs115982800 was associated with age at baseline. Points to the left of the vertical line would suggest the variant was associated with a younger age at baseline.

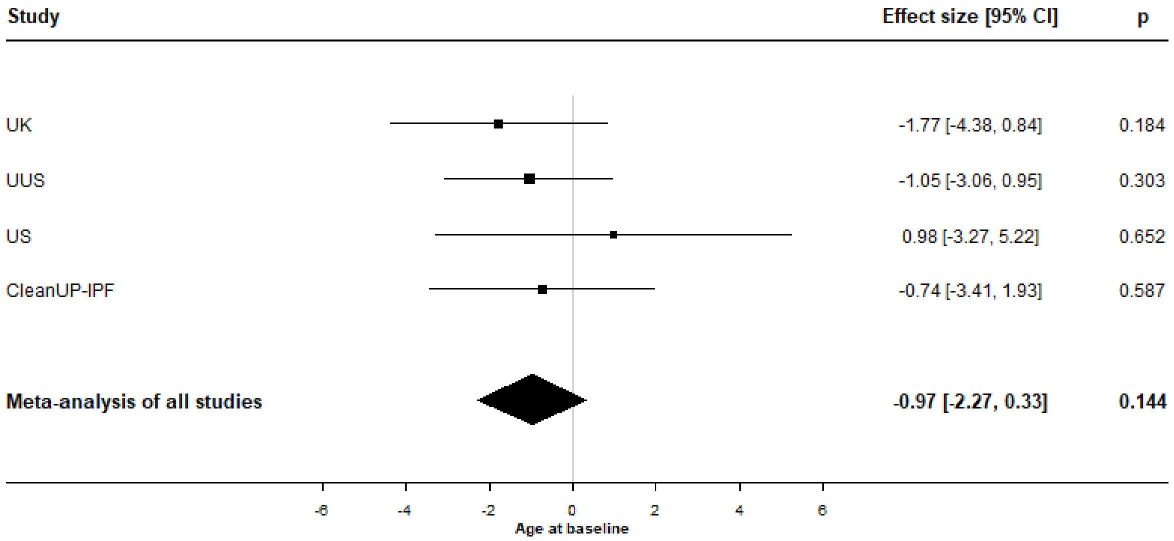

**vi) Survival model**

Results when testing the association of rs115982800 with transplant-free survival times after diagnosis of IPF. Points to the right of the vertical line would suggest the variant was associated with increased mortality. Results are given as hazard ratios and plotted on a log scale. As there were no deaths recorded for individuals in the CleanUP-IPF study with a copy of the A allele, the standard errors were very large.

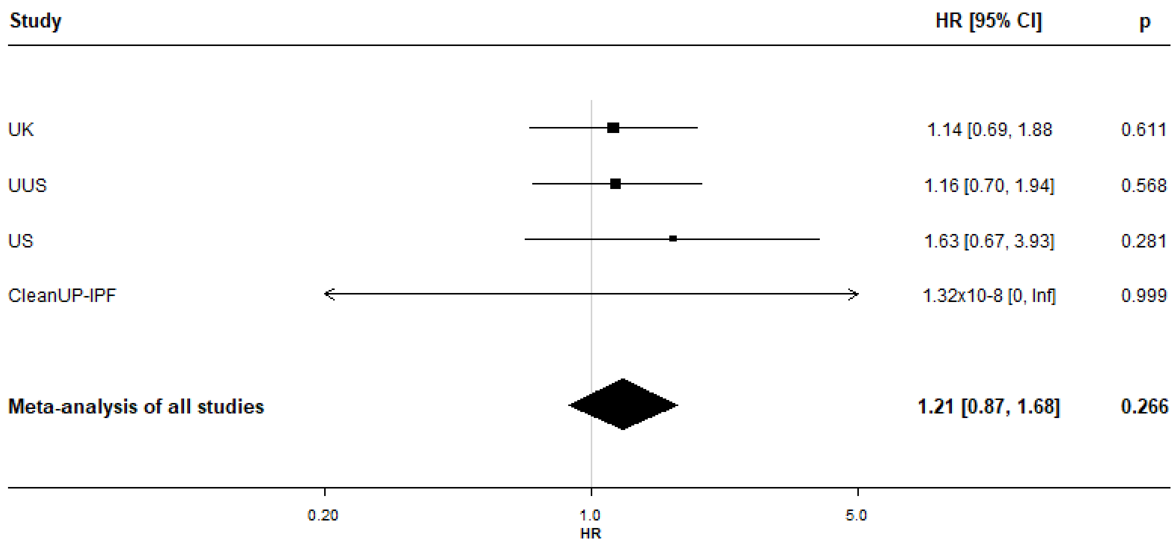

### Supplementary Figure 8: Kaplan-Meier plots split by rs115982800 genotype

Kaplan-Meier plots for transplant-free survival times. The x axis shows time with the estimated survival function S(t) on the y axis. Individuals with a GG genotype are shown in black, individuals with a GA genotype are shown in red and those with an AA genotype are shown in blue. As can be seen in the fourth plot, no individuals with an rs115982800_A allele (red and blue lines) had a recorded event in the CleanUP study.

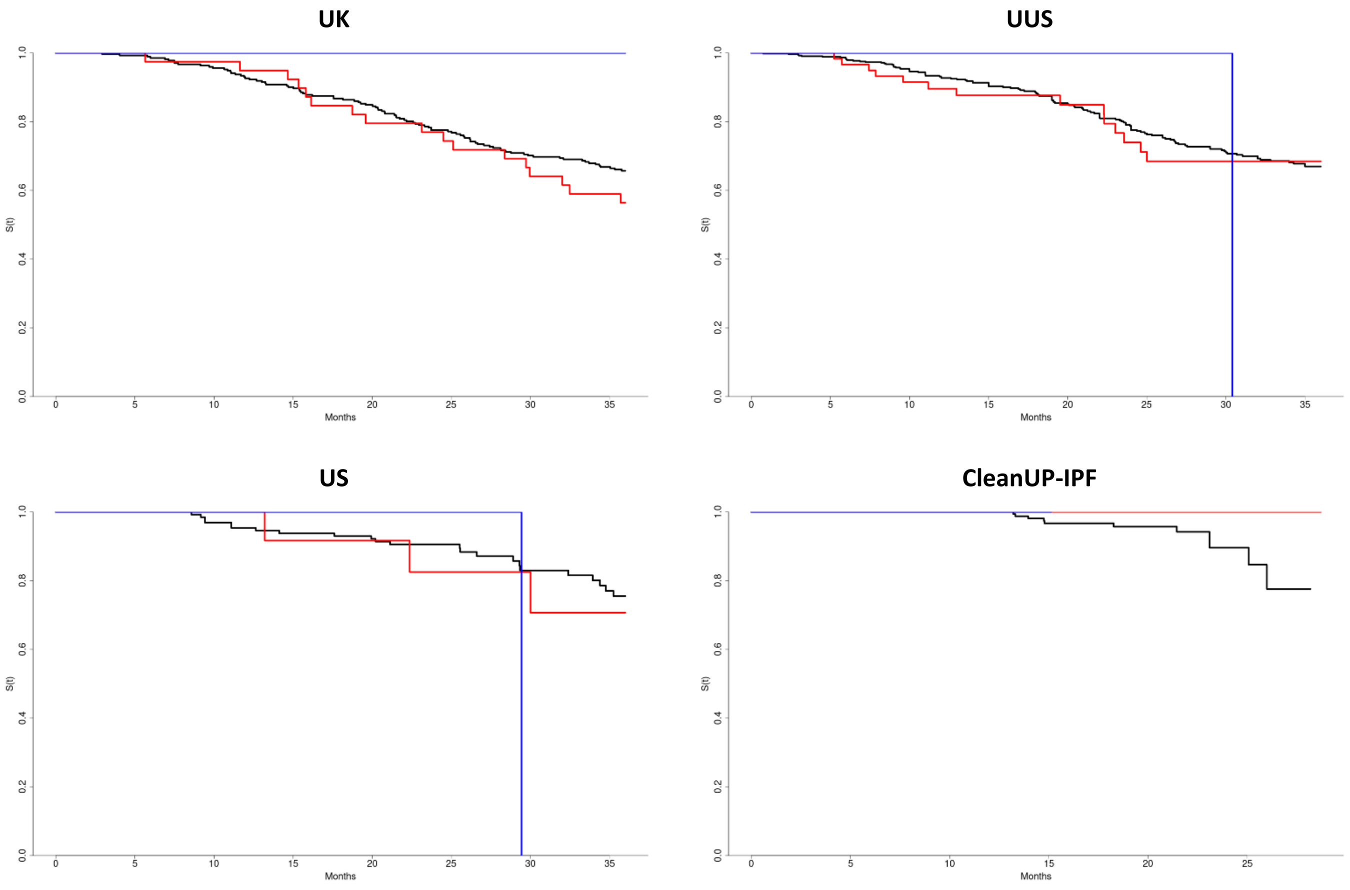

**Supplementary Figure 9: Summary of Hi-C results**

The plot below summarises the Hi-C results for testing whether the fragment containing rs115982800 (shown by the solid red bar) physically interacts with any other fragments in the region (within 3Mb). Each white or grey background bar denotes the location of each DNA fragment with position on the x axis. Only physical interactions that were significant after Bonferroni correction for that tissue are plotted (–log(p value) on the y axis). Points are coloured by the tissue that the association was observed in (a fragment can show an association in multiple tissues). The green boxes at the top show the location of nearby genes that are located (or partially located) in a fragment that shows a significant interaction with the fragment containing rs115982800.

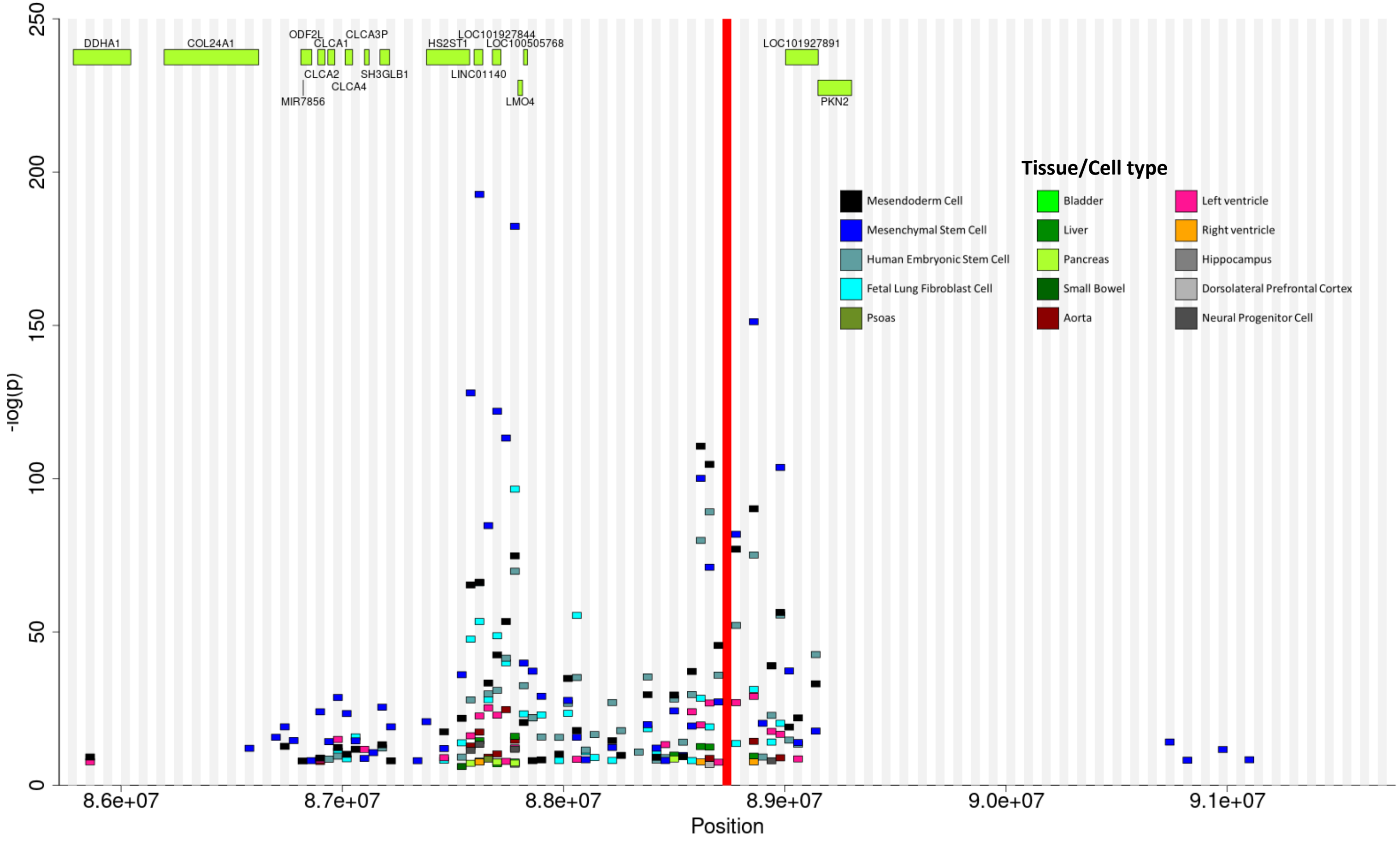

**Supplementary Figure 10: Mirror plot comparing longitudinal FVC association signal with *CCBL2* gene expression**

Each point represents a genetic variant with position on the x axis and the –log(p value) on the y axis. Points above the x axis show the association of the variant with longitudinal FVC after diagnosis of IPF and points below the axis show the association of the variant with *CCBL2* gene expression in whole blood (eQTLGen). Points are coloured by LD with the longitudinal FVC variant rs115982800 (shown in blue); variants with $r^2 \geq 0.8$ are shown in red, variants with $0.8 > r^2 \geq 0.6$ in orange, variants with $0.6 > r^2 \geq 0.4$ in yellow, variants with $0.4 > r^2 \geq 0.4$ in light yellow and variants with $r^2 < 0.2$ in grey. The red horizontal line shows the genome-wide significance threshold ($p = 5 \times 10^{-8}$). The location of the *CCBL2* gene is shown by the green box. Plots were generated using the "mirrorplot" R package (https://github.com/rjallen513/mirrorplot).

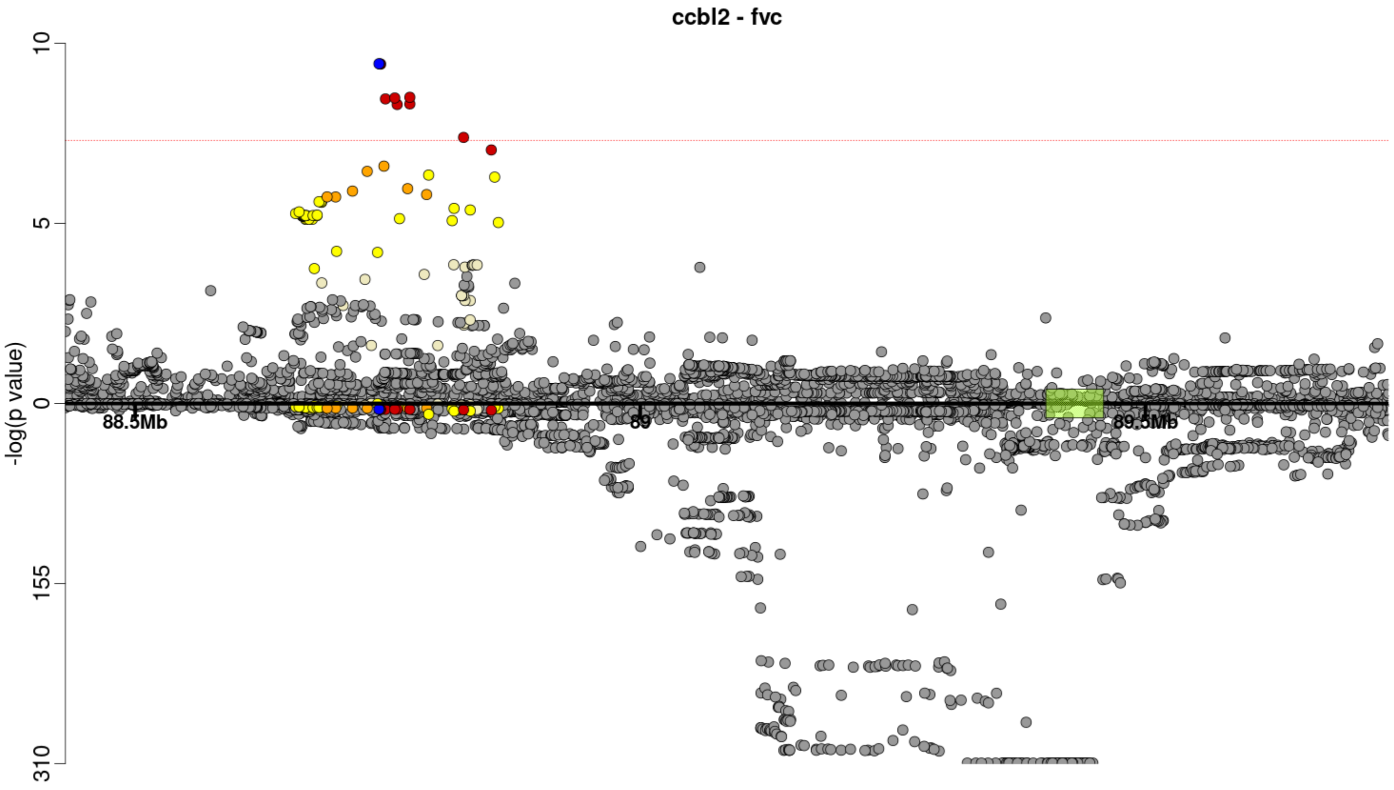

**Supplementary Figure 11: Summary of gene prioritisation analyses.**

This table summarises where there is evidence to support the association signal acting through the given gene. Red squares show where there is evidence to support the gene, orange shows where there is weak or inconclusive evidence to support the gene.

| Chr | Start | End | Gene | Nearest gene | Annotation | Gene expression | Physical interactions | Mendelian diseases | Rare variant PheWAS | Mouse knockouts |
|---|---|---|---|---|---|---|---|---|---|---|
| 1 | 85715636 | 85725355 | C1orf52 | | | | | | | |
| 1 | 85731459 | 85742587 | *BCL10* | | | | | | | |
| 1 | 85742040 | 85743771 | *LOC646626* | | | | | | | |
| 1 | 85784167 | 86044046 | *DDAH1* | | | | | | | |
| 1 | 86046443 | 86049648 | *CYR61* | | | | | | | |
| 1 | 86115105 | 86174116 | *ZNHIT6* | | | | | | | |
| 1 | 86194915 | 86622121 | *COL24A1* | | | | | | | |
| 1 | 86812506 | 86862025 | *ODF2L* | | | | | | | |
| 1 | 86823314 | 86823370 | *MIR7856* | | | | | | | |
| 1 | 86889768 | 86922240 | *CLCA2* | | | | | | | |
| 1 | 86934525 | 86965974 | *CLCA1* | | | | | | | |
| 1 | 87012758 | 87046432 | *CLCA4* | | | | | | | |
| 1 | 87099958 | 87121059 | *CLCA3P* | | | | | | | |
| 1 | 87170252 | 87213867 | *SH3GLB1* | | | | | | | |
| 1 | 87328127 | 87380107 | *SEP15* | | | | | | | |
| 1 | 87380334 | 87575681 | *HS2ST1* | | | | | | | |
| 1 | 87595447 | 87634886 | *LINC01140* | | | | | | | |
| 1 | 87678351 | 87717014 | *LOC101927844* | | | | | | | |
| 1 | 87794150 | 87814607 | *LMO4* | | | | | | | |
| 1 | 87819209 | 87837338 | *LOC100505768* | | | | | | | |
| 1 | 89003195 | 89150887 | *LOC101927891* | | | | | | | |
| 1 | 89149921 | 89301938 | *PKN2* | | | | | | | |
| 1 | 89318320 | 89357301 | *GTF2B* | | | | | | | |
| 1 | 89401455 | 89458643 | *CCBL2* | | | | | | | |
| 1 | 89445138 | 89458643 | *RBMXL1* | | | | | | | |
| 1 | 89472359 | 89488549 | *GBP3* | | | | | | | |
| 1 | 89517986 | 89531043 | *GBP1* | | | | | | | |
| 1 | 89571815 | 89591842 | *GBP2* | | | | | | | |
| 1 | 89597433 | 89641723 | *GBP7* | | | | | | | |
| 1 | 89646830 | 89664633 | *GBP4* | | | | | | | |
| 1 | 89724633 | 89738544 | *GBP5* | | | | | | | |
| 1 | 89754940 | 89756045 | *LOC729930* | | | | | | | |
| 1 | 89829435 | 89853719 | *GBP6* | | | | | | | |
| 1 | 89873237 | 89890493 | *GBP1P1* | | | | | | | |
| 1 | 89990396 | 90063420 | *LRRC8B* | | | | | | | |
| 1 | 90090407 | 90098453 | *FLJ27354* | | | | | | | |
| 1 | 90098643 | 90185094 | *LRRC8C* | | | | | | | |
| 1 | 90286572 | 90401989 | *LRRC8D* | | | | | | | |
| 1 | 90458823 | 90460525 | *GEMIN8P4* | | | | | | | |
| 1 | 90460677 | 90494094 | *ZNF326* | | | | | | | |
| 1 | 91177578 | 91182794 | *BARHL2* | | | | | | | |
| 1 | 91380856 | 91487812 | *ZNF644* | | | | | | | |
| 1 | 91726322 | 91870426 | *HFM1* | | | | | | | |

### Supplementary Figure 12: Manhattan plots for gene-based analyses

Each point represents a gene with midpoint chromosomal position on the x axis and –log(p value) for association with FVC or DLco decline on the y axis. The red line shows the significance threshold used.

#### i) FVC

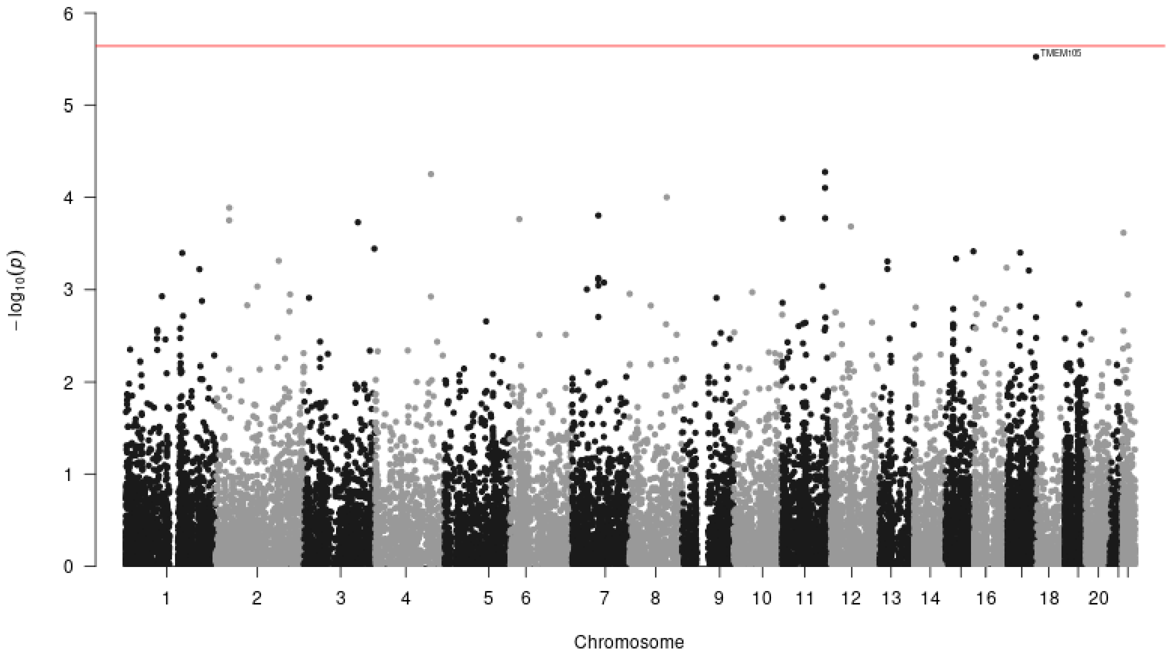

#### ii) DLco

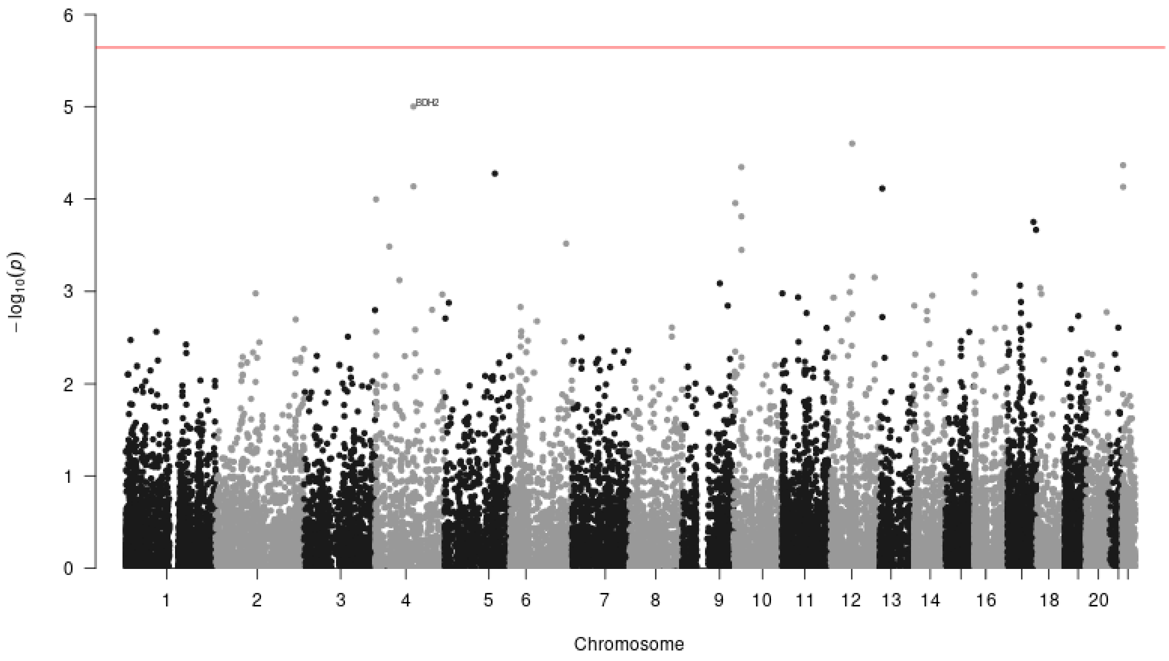
